# Towards achieving a vaccine-derived herd immunity threshold for COVID-19 in the U.S.

**DOI:** 10.1101/2020.12.11.20247916

**Authors:** Abba B. Gumel, Enahoro A. Iboi, Calistus N. Ngonghala, Gideon A. Ngwa

## Abstract

A novel coronavirus emerged in December of 2019 (COVID-19), causing a pandemic that continues to inflict unprecedented public health and economic burden in all nooks and corners of the world. Although the control of COVID-19 has largely focused on the use of basic public health measures (primarily based on using non-pharmaceutical interventions, such as quarantine, isolation, social-distancing, face mask usage and community lockdowns), three safe and highly-effective vaccines (by AstraZeneca Inc., Moderna Inc. and Pfizer Inc., with protective efficacy of 70%, 94.1% and 95%, respectively) have been approved for use in humans since December 2020. We present a new mathematical model for assessing the population-level impact of the three currently-available anti-COVID vaccines that are administered in humans. The model stratifies the total population into two subgroups, based on whether or not they habitually wear face mask in public. The resulting multigroup model, which takes the form of a deterministic system of nonlinear differential equations, is fitted and parametrized using COVID-19 cumulative mortality data for the third wave of the COVID-19 pandemic in the U.S. Conditions for the asymptotic stability of the associated disease-free equilibrium, as well as expression for the vaccine-derived herd immunity threshold, are rigorously derived. Numerical simulations of the model show that the size of the initial proportion of individuals in the masks-wearing group, together with positive change in behaviour from the non-masks wearing group (as well as those in masks-wearing group do not abandon their masks-wearing habit) play a crucial role in effectively curtailing the COVID-19 pandemic in the U.S. This study further shows that the prospect of achieving herd immunity (required for COVID-19 elimination) in the U.S., using any of the three currently-available vaccines, is quite promising. In particular, while the use of the AstraZeneca vaccine will lead to herd immunity in the U.S. if at least 80% of the populace is vaccinated, such herd immunity can be achieved using either the Moderna or Pfizer vaccine if about 60% of the U.S. population is vaccinated. Furthermore, the prospect of eliminating the pandemic in the US in the year 2021 is significantly enhanced if the vaccination program is complemented with nonpharmaceutical interventions at moderate increased levels of compliance (in relation to their baseline compliance). The study further suggests that, while the waning of natural and vaccine-derived immunity against COVID-19 induces only a marginal increase in the burden and projected time-to-elimination of the pandemic, adding the impacts of the therapeutic benefits of the vaccines into the model resulted in a dramatic reduction in the burden and time-to-elimination of the pandemic.

## 1 Introduction

The novel coronavirus (COVID-19) pandemic, which started as a pneumonia of an unknown etiology late in December 2019 in the city of Wuhan, became the most devastating public health challenge mankind has faced since the 1918/1919 pandemic of influenza. The COVID-19 pandemic, which rapidly spread to essentially every nook and corner of the planet, continues to inflict devastating public health and economic challenges globally. As of January 24, 2021, the pandemic accounted for about 100 million confirmed cases and 2, 128, 721 cumulative mortality globally. Similarly, as of this date, the United States, which recorded its first COVID-19 case on January 20, 2020, recorded over 25, 123, 857 confirmed cases and 419, 204 deaths [1].

COVID-19, a member of the Coronavirus family of RNA viruses is primarily transmitted from human-to-human through inhalation of respiratory droplets from both symptomatic and asymptomatically-infectious humans [2] (*albeit* there is limited evidence that COVID-19 can be transmitted *via* exhalation through normal breathing and aerosol [3]. The incubation period of the disease is estimated to lie between 2 to 14 days (with a mean of 5.1 days), and majority of individuals infected with the disease show mild or no clinical symptoms [4]. The symptoms typically include coughing, fever and shortness of breadth (for mild cases) and pneumonia for severe cases [4]. The people most at risk of dying from, or suffering severe illness with, COVID-19 are those with co-morbidities (such as individuals with diabetes, obesity, kidney disease, cardiovascular disease, chronic respiratory disease, etc.). Younger people, front line healthcare workers and employees who maintain close contacts (within 6 feet) with customers and other co-workers (such as meat factory workers, retail store workers, etc.) are also at risk.

Prior to the approval of the three safe and effective vaccines (by AstraZeneca, Moderna and Pfizer) for use in humans in December 2020 [5, 6], the control and mitigation efforts against COVID-19 have been focused on the use of non-pharmaceutical interventions (NPIs), such as quarantine, self-isolation, social (physical) distancing, the use of face masks in public, hand washing (with approved sanitizers), community lockdowns, testing and contact tracing. Of these NPIs, the use of face masks in public was considered to be the main mechanism for effectively curtailing COVID-19 [4, 7–9]. Furthermore, owing to its limited supply, the approved anti-COVID drug *remdesivir* is reserved for use to treat individuals in hospital who display severe symptoms of COVID-19. The US started administering the Pfizer and Moderna vaccines by December 2020 [5, 6].

The Pfizer and Moderna vaccines, each offering a protective efficacy of about 95% [10–12], are genetic vaccines that trigger the immune system to recognize the coronavirus’ spike protein and develop antibodies against it [10, 13]. Two doses are required for both the Pfizer and Moderna vaccines (one to prime the immune system, and the second to boost it). For the Pfizer vaccine, the second dose is administered 19-42 days after the first dose, while that for the Moderna vaccine is administered three to four weeks after the first dose. Both vaccines need to be stored at appropriate refrigeration temperatures [14]. The AstraZeneca vaccine, on the other hand, has estimated protective efficacy of 70% [10–12]. It uses a replication-deficient chimpanzee viral vector that contains the genetic material of the SARS-CoV-2 virus spike protein [12]. The AstraZeneca vaccine also requires two doses (one month apart) to achieve immunity, and unlike the Pfizer and Moderna vaccines, does not have to be stored in super-cold temperatures [12].

A vaccine, when effective, can offer different levels of protection to the vaccinated person, with the protection ranging from reducing or blocking probability of acquiring infection (for very effective vaccines) to reduction of severity of disease, hospitalization, and mortality and accelerating recovery in breakthrough infections (for vaccines that offer strong therapeutic benefits) [15, 16]. A vaccine that has protective ability, when introduced into a population during an epidemic, will have an important consequence on the progression of the epidemic, and its effective deployment would be dependent on the strategy used. Optimal vaccination outcomes can be achieved if the vaccination programs are well-conceived and monitored. In the absence of empirical data during the epidemic, mathematical models can offer a plausible pathway to predicting the effectiveness of targeted vaccination programs. The goal of this study is to design a structured mathematical model that will allow for the realistic assessment of the population-level impact of vaccination programs based on using the three vaccines, with emphasis on determining the optimal coverage rate needed to achieve vaccine-derived *herd immunity* (which is required for eliminating the pandemic). A secondary objective is to explore whether the prospect for eliminating the pandemic in the US will be enhanced if the vaccination program is combined with NPIs, such as social-distancing at some level of compliance.

Numerous mathematical models, of various types, have been developed and used to provide insight into the transmission dynamics and control of COVID-19. The modeling types used include statistical [17], compartmental/deterministic (e.g., [4, 7–9, 18–20]), stochastic (e.g., [21, 22]), network (e.g., [23]) and agent-based (e.g., [24]). A notable feature of the model to be developed in this project is its multigroup nature. Specifically, the total population will be subdivided into two groups, namely those who habitually wear face mask in public and those who do not. Cumulative mortality data for COVID-19 pandemic in the U.S. will be used to parametrize the model. The expected outcome of the study is the determination of the minimum vaccine coverage level needed to effectively curtail (or eliminate) community transmission of COVID-19 in the U.S., and quantify the reduction in the required vaccine coverage if the vaccination program is supplemented with face masks usage (under various face masks efficacy and compliance parameter space). The rest of the paper is organized as follows. The novel multigroup model is formulated in Section 2. The parameters of the model are also estimated, based on fitting the model with U.S. COVID-19 mortality data for the third wave of the pandemic. The model is rigorously analysed, with respect to the asymptotic stability of the disease-free equilibrium of the model, in Section 3. A condition for achieving community-wide vaccine-derived herd immunity is also derived. Numerical simulations of the model are reported in Section 4. Discussions and concluding remarks are presented in Section 5.

## 2 Formulation of Mathematical Model

In order to account for heterogeneity in face masks usage in the community, the total population of individuals in the community at time *t*, denoted by *N* (*t*), is split into the total sub-populations of individuals who do not habitually wear face mask in public (labeled “*non-mask users*”), denoted by *N*_1_(*t*), and the total sub-populations of those who habitually wear face mask in public (labeled “*mask users*”), represented by *N*_2_(*t*). That is, *N* (*t*) = *N*_1_(*t*) + *N*_2_(*t*). Furthermore, the sub-population *N*_1_(*t*) is sub-divided into the mutually-exclusive compartments of unvaccinated susceptible (*S*_1*u*_(*t*)), vaccinated susceptible (*S*_1*v*_(*t*)), exposed (*E*_1_(*t*)), pre-symptomatically-infectious (*P*_1_(*t*)), symptomatically-infectious (*I*_1_(*t*)), asymptomatically-infectious (*A*_1_(*t*)), hospitalized (*H*_1_(*t*)) and recovered (*R*_1_(*t*)) individuals, so that

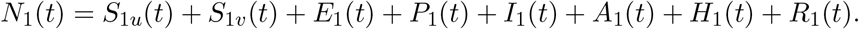

Similarly, the total sub-population of the mask users, *N*_2_(*t*), is stratified into the compartments for unvaccinated susceptible (*S*_2*u*_(*t*)), vaccinated susceptible (*S*_2*v*_(*t*)), exposed (*E*_2_(*t*)), pre-symptomatically-infectious (*P*_2_(*t*)), symptomatically-infectious (*I*_2_(*t*)), asymptomatically-infectious (*A*_2_(*t*)), hospitalized (*H*_2_(*t*)) and recovered (*R*_2_(*t*)) individuals. Hence,

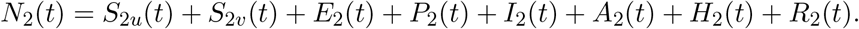

### 2.1 Infection Rates

In this section, the functional form of the infection rate (or effective contact rate) for a susceptible individual in group 1 or 2 will be derived. The model to be formulated has four infectious classes, namely the classes for presymptomatic (*P*_*i*_), symptomatic (*I*_*i*_), asymptomatic (*A*_*i*_) and hospitalized (*H*_*i*_) individuals (*i* = 1, 2). Hence, the rate at which an individual in group *i* acquires infection from an infectious individual in any of the four infectious classes is given by the average number of contacts *per* unit time (measured in days) for susceptible individuals (denoted by *c*_*k*_; with *k* = {*P*_*i*_, *I*_*i*_, *A*_*i*_, *H*_*i*_*}*and *i* = 1, 2), *times* the sum (over all infectious compartments in group *i*) of the probability of transmission *per* contact with an infectious individual in group *i* (denoted by 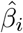) *times* the probability that a random infectious contact the susceptible individual makes is with an infectious individual in group *i* (denoted by *ρ*_*k*_).

Let *c*_*k*_ be the average number of contacts an individual in epidemiological compartment *k* makes *per* unit time. It then follows that the probability that a random contact this individual makes is with someone else in epidemiological compartment *k* is given by the total number contacts made by everyone in that compartment, denoted by *c*_*ki*_, divided by the total number of contacts for the entire population. That is, 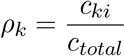, where

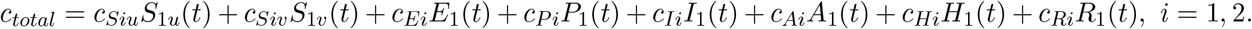

Based on the above definitions, it follows that the infection rate of a susceptible individual in group *i*, denoted by *λ*_*i*_ (*i* = 1, 2), is given by

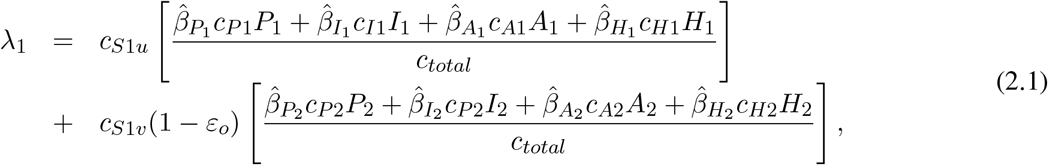

Similarly, the infection rate for a susceptible individual in group 2, denoted by *λ*_2_, is given by:

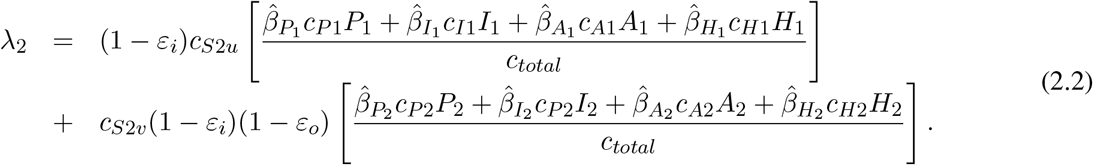

In (2.1) and (2.2), the parameters 0 *< ε*_*o*_ *<* 1 and 0 *< ε*_*i*_ *<* 1 represent the outward and inward protective efficacy, respectively, of face masks to prevent the transmission of infection to a susceptible individual (*ε*_*o*_) as well as prevent the acquisition of infection (*ε*_*i*_) from an infectious individual. For mathematical tractability (needed to reduce the number of parameters of the model to be developed), we assume that every member of the population has the same number of contacts. That is, we assume that *c*_*S*1*u*_ = *c*_*S*1*v*_ =… = *c*_*R*2_ = *k*_*c*_. Hence, *c*_*total*_ = *k*_*c*_*N* (*t*). Let 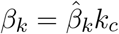. Using this definition of *β*_*k*_ and *c*_*total*_ = *k*_*c*_*N* in Equations (2.1) and (2.2) gives, respectively,

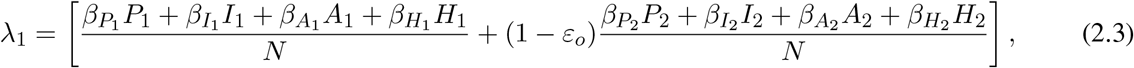

and,

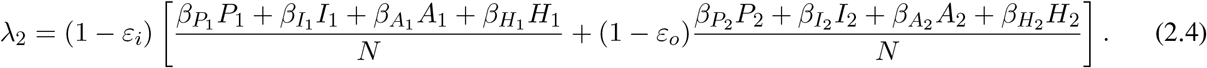

### 2.2 Equations of Mathematical Model

Before giving the equations for the two-group vaccination model, it is important to recall that vaccination against COVID-19 in the US is administered to individuals of a certain eligible age (e.g., 12 years of age and older for the Pfizer vaccine and 18 years of age and older for the Moderna vaccine). Consequently, in formulating a model that incorporates COVID-19 vaccines, it is important that demographic parameters (birth and natural death) are included to account for the new cohort of susceptible individuals that reach the minimum eligible age for receiving the vaccine. The equations for the rate of change of the sub-populations of non-mask users (i.e., individuals in group 1) is given by the following deterministic system of nonlinear differential equations (where a dot represents differentiation with respect to time *t*):

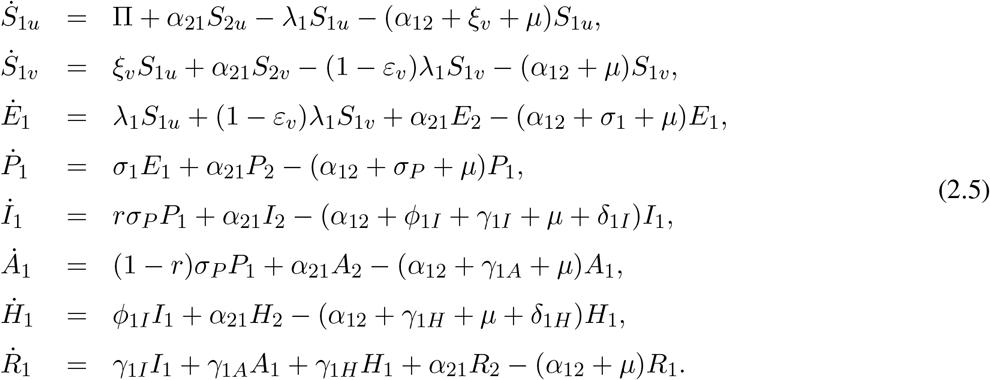

where, *λ*_1_ is as defined in (2.3).

In Equation (2.5), the parameter Π is the recruitment rate into the population (this parameter also captures the inflow of new susceptible individuals that have reached the minimum eligibility age for getting a vaccine). Furthermore, *α*_21_ is the rate at which individuals in the habitual mask-wearing group 2 change their behavior and move to the non-masking group 1, and *α*_12_ is the rate at which individuals in group 1 change their nonmasking behavior and move to group 2. For mathematical tractability, we do not distinguish the change of behavior parameters (*α*_12_ and *α*_21_) for unvaccinated and vaccinated individuals, and we assume that all recruited individuals (at the rate Π) are initially in the non-masking group. The parameter *ξ*_*v*_ represents the *per capita* vaccination rate, and the vaccine is assumed to induce protective efficacy 0 *< ε*_*v*_ *<* 1 in all vaccinated individuals (i.e., the vaccine is imperfect). Natural deaths occurs in all epidemiological classes at a rate *µ*. Individuals in the *E*_1_ class progress to the pre-symptomatic stage at a rate *σ*_1_, and those in the pre-symptomatic class (*P*_1_) transition out of this class at a rate *σ*_*P*_ (a proportion, *q*, of which become symptomatic, and move to the *I* class at a rate *qσ*_*P*_, and the remaining proportion, 1 − *q*, move to the asymptomatically-infectious class at a rate (1 − *q*)*σ*_*P*_). Symptomatic infectious individuals are hospitalized at a rate *ϕ*_1*I*_. They recover at a rate *γ*_1*I*_ and die due to the disease at a rate *δ*_1*I*_. Hospitalized individuals die of the disease at the rate *δ*_1*H*_.

Similarly, the equations for the rate of change of the sub-populations of mask users (i.e., individuals in group 2) is given by the following system of nonlinear differential equations:

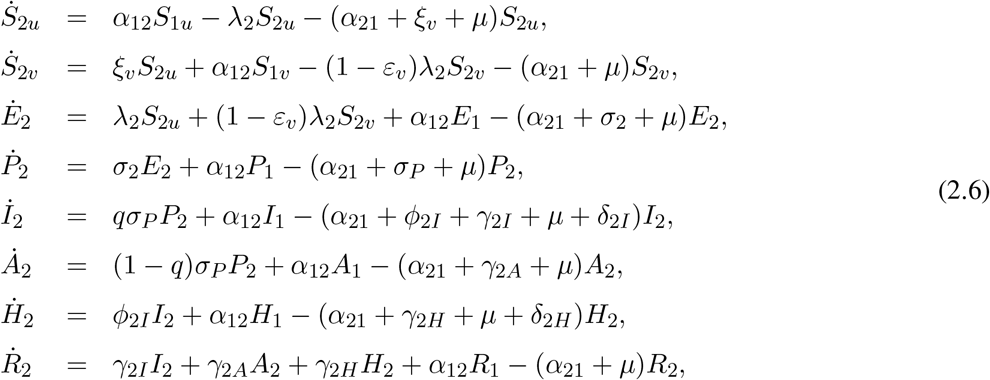

with *λ*_2_ defined in (2.4). Thus, Equations (2.5) and (2.6) represent the multi-group model for assessing the population impact of face masks usage and vaccination on the transmission dynamics and control of COVID-19 in a community. The flow diagram of the model {(2.5), (2.6)} is depicted in Figure 1 (the state variables and parameters of the model are described in Tables 1 and 2, respectively).

**Figure 1:**
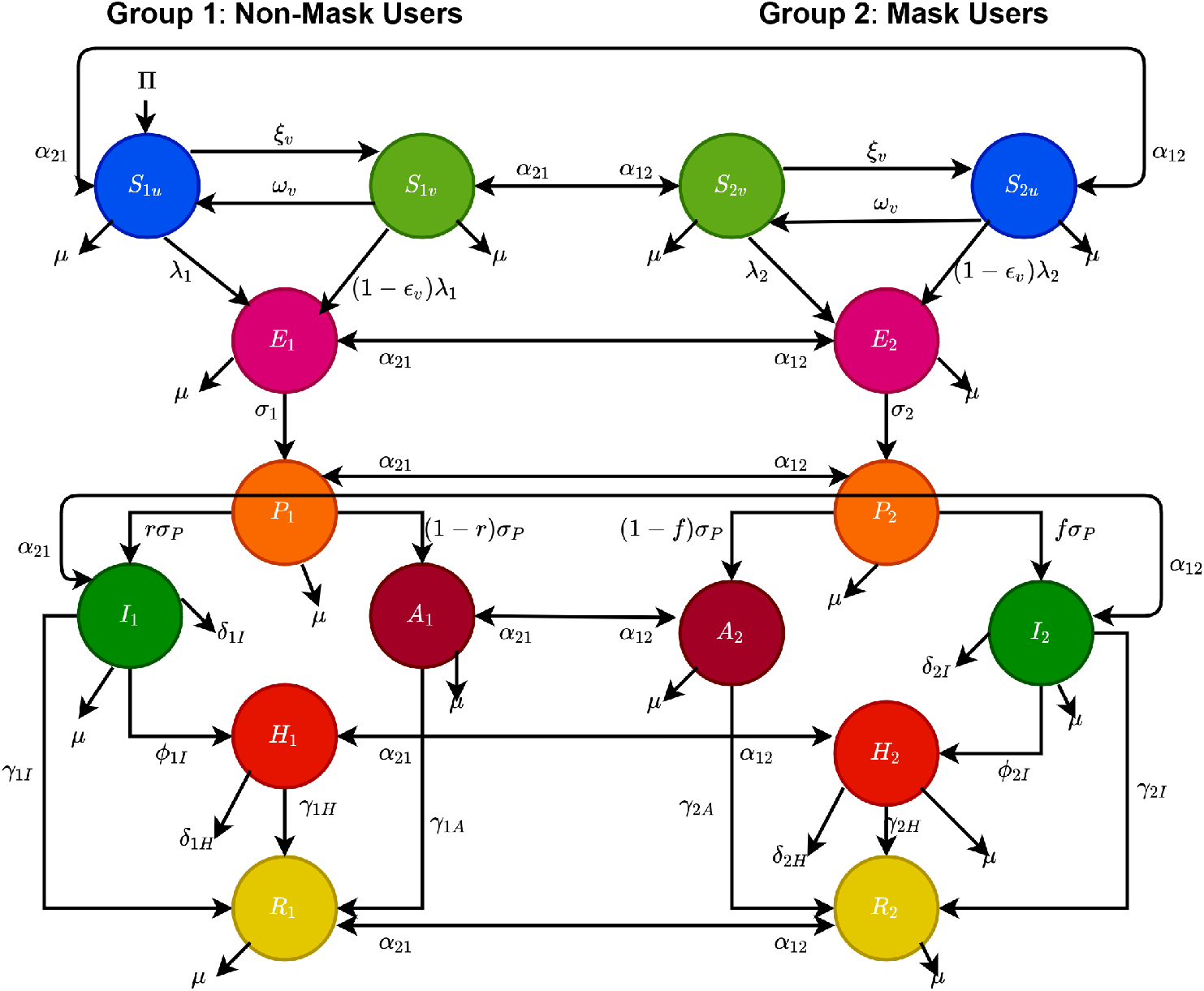
Flow diagram of the model {(2.5), (2.6)}.

**Table 1:** Description of the state variables of the model {(2.5), (2.6)}.

| State variable | Description |
| --- | --- |
| $S_{1u}$ | Population of non-vaccinated susceptible individuals who do not habitually wear face masks |
| $S_{2u}$ | Population of non-vaccinated susceptible individuals who habitually face masks |
| $S_{1v}$ | Population of vaccinated susceptible individuals who do not habitually wear face masks |
| $S_{2v}$ | Population of vaccinated susceptible individuals who habitually wear face masks |
| $E_1$ | Population of exposed (newly-infected) individuals who do not habitually wear face masks |
| $E_2$ | Population of exposed (newly-infected) individuals who habitually wear face masks |
| $P_1$ | Population of pre-symptomatic infectious individuals who do not habitually wear face masks |
| $P_2$ | Population of pre-symptomatic infectious individuals who habitually wear face masks |
| $I_1$ | Population of symptomatically-infectious individuals who do not habitually wear face masks |
| $I_2$ | Population of symptomatically-infectious individuals who habitually wear face masks |
| $A_1$ | Population of asymptotically-infectious individuals who do not habitually wear face masks |
| $A_2$ | Population of asymptotically-infectious individuals who habitually wear face masks |
| $H_1$ | Population of hospitalized individuals who do not habitually wear face masks |
| $H_2$ | Population of hospitalized individuals who habitually wear face masks |
| $R_1$ | Population of recovered individuals who do not habitually wear face masks |
| $R_2$ | Population of recovered individuals who habitually wear face masks |

**Table 2:** Description of the parameters of the model {(2.5), (2.6)}.

| Parameters | Description |
| --- | --- |
| $\Pi$ | Recruitment rate into the population |
| $\mu$ | Natural mortality rate |
| $\beta_{P1}(\beta_{P2})$ | Effective contact rate for pre-symptomatic individuals who do not wear (wear) face masks |
| $\beta_{I1}(\beta_{I2})$ | Effective contact rate for infectious symptomatic individuals who do not wear (wear) face masks |
| $\beta_{A1}(\beta_{A2})$ | Effective contact rate for symptomatically-infectious individuals who do not wear (wear) face masks |
| $\beta_{H1}(\beta_{H2})$ | Effective contact rate for hospitalized individuals who do not wear (wear) face masks |
| $0 < \epsilon_0 < 1$ | Outward protective efficacy of face masks |
| $0 < \epsilon_i < 1$ | Inward protective efficacy of face masks |
| $\alpha_{12}$ | Rate at which non-habitual face masks wearers choose to become habitual wearers |
| $\alpha_{21}$ | Rate at which habitual face masks wearers choose to become non-habitual wearers |
| $\xi_v$ | *Per capita* vaccination rate |
| $0 < \varepsilon_v < 1$ | Protective efficacy of the vaccine |
| $\sigma_1(\sigma_2)$ | Rate at which exposed individuals who do not wear (wear) face masks progress to the corresponding pre-symptomatic infectious stage |
| $\sigma_P$ | Rate at which pre-symptomatic infectious individuals progress to symptomatically-infectious or asymptotically-infectious stage |
| $r(q)$ | Proportion of pre-symptomatic infectious individuals who do not wear (wear) face masks that become symptomatically-infectious |
| $\phi_{1I}(\phi_{2I})$ | Hospitalization rate for symptomatically-infectious individuals who do not wear (wear) face masks |
| $\gamma_{1A}(\gamma_{2A})$ | Recovery rate for asymptotically-infectious individuals who do not wear (wear) face masks |
| $\gamma_{1I}(\gamma_{2I})$ | Recovery rate for symptomatically-infectious individuals who do not wear (wear) face masks |
| $\gamma_{1H}(\gamma_{2H})$ | Recovery rate for hospitalized individuals who do not wear (wear) face masks |
| $\delta_{1I}(\delta_{2I})$ | Disease-induced mortality rate for symptomatically-infectious individuals who do not wear (wear) face masks |
| $\delta_{1H}(\delta_{2H})$ | Disease-induced mortality rate for hospitalized individuals who do not wear (wear) face masks |

The multi-group model {(2.5), (2.6)} is an extension of the two-group mask-use model in [7] by, *inter alia*:

i. allowing for back-and-forth transitions between the two groups (mask-users and non-mask-users), to account for human behavioral changes *vis a vis* decision to either be (or not to be) a face mask user in public;
ii. incorporating an imperfect vaccine, which offers protective efficacy (0 *< ε*_*v*_ *<* 1) against acquisition of COVID-19 infection;
iii. allowing for disease transmission by pre-symptomatic and asymptomatically-infectious individuals.

### 2.3 Data Fitting and Parameter Estimation

In this section, cumulative COVID-19 mortality data for the U.S. (for the period October 12, 2020 to January 20, 2021) will be used to fit the model (2.5)-(2.6) in the absence of vaccination. The fitting will allow us to estimate some of the key (unknown) parameters of the model. In particular, the parameters to be estimated from the data are the community transmission rate for individuals who do not wear face masks in public (*β*_1_), the transmission rate for individuals who habitually wear face masks in public (*β*_2_), the inward efficacy of masks in preventing disease acquisition by susceptible individuals who habitually wear face masks (*ε*_*i*_), the outward efficacy of masks to prevent the spread of disease by infected individuals who habitually wear face masks (*ε*_*o*_), the rate at which people who do not wear masks adopt a mask-wearing habit (*α*_12_), the rate at which those who habitually wear face masks stop wearing masks in public (*α*_21_), and the mortality rates of symptomatic infectious and hospitalized individuals (*δ*_*i*_ and *δ*_*h*_, respectively). It should be mentioned that modification parameters *η*_*P*_, *η*_*I*_, *η*_*A*_, and *η*_*H*_ relating to disease transmission by pre-symptomatic infectious, symptomatic infectious, asymptomatic infectious and hospitalized individuals, respectively, are introduced in the forces of infection *λ*_1_ and *λ*_2_, so that *β*_*j*_ = *η*_*j*_*β*_*k*_ (*j* ∈{*P*_*k*_, *I*_*k*_, *A*_*k*_, *H*_*k*_*}, k* ∈{1, 2}). The model was fitted using a standard nonlinear least squares approach, which involved using the inbuilt MATLAB minimization function *“lsqcurvefit”* to minimize the sum of the squared differences between each observed cumulative mortality data point and the corresponding cumulative mortality point obtained from the model (2.5)-(2.6) in the absence of vaccination [4, 25, 26]. The choice of mortality over case data is motivated by the fact that mortality data for COVID-19 is more reliable than case data (see [8] for details). The estimated values of the fitted parameters, together with their 95% confidence intervals, are tabulated in Table 3. The (fixed) values of the remaining parameters of the model are tabulated in Table 4. Figure 2 depicts the fitting of the model to the observed cumulative COVID-19 mortality data for the U.S. Furthermore, Figure 2 compares the simulations of the model using the fitted (estimated) and fixed parameters (given in Tables 3 and 4) with the observed daily COVID-19 mortality for the US.

**Table 3:** Estimated (fitted) parameter values and their 95% confidence intervals for the model (2.5)-(2.6) in the absence of vaccination, using COVID-19 mortality data for the US for the period from October 12, 2020 January 20, 2021.

| Parameter | Value | Confidence interval |
| --- | --- | --- |
| $\beta_1$ | 0.224334/day | [0.201828, 0.370926]/day |
| $\beta_2$ | 0.072957/day | [0.000002, 0.178140]/day |
| $\varepsilon_o$ | 0.507666 (dimensionless) | [0.282518, 0.692441] (dimensionless) |
| $\varepsilon_i$ | 0.623667 (dimensionless) | [0.020807, 0.999999] (dimensionless) |
| $\alpha_{12}$ | 0.006229/day | [0.004732, 0.008508]/day |
| $\alpha_{21}$ | 0.000798/day | [0.000000, 0.000999]/day |
| $\delta_i$ | 0.000573/day | [0.000000, 0.002399]/day |
| $\delta_h$ | 0.009505/day | [0.000708, 0.011030]/day |

**Table 4:** Baseline values of the fixed parameters of the model (2.5)-(2.6).

| Parameter | Value | Source | Parameter | Value | Source |
| --- | --- | --- | --- | --- | --- |
| $\sigma_1 (\sigma_2)$ | 1/2.5 (1/2.5)/day | [27, 28] | $\Pi$ | $1.2 \times 10^4$/day | Estimated |
| $\sigma_p$ | 1/2.5/day | [27, 28] | $\mu$ | 1/(79 × 365)/day | Estimated |
| $r (q)$ | 0.2(0.2) (dimensionless) | [29, 30] | $\eta_P$ | 1.25 (dimensionless) | Assumed |
| $\phi_{1I}$ | 1/6/day | [31] | $\eta_I$ | 1.0 (dimensionless) | Assumed |
| $\phi_{2I}$ | 1/6/day | [31] | $\eta_A$ | 1.50 (dimensionless) | Assumed |
| $\gamma_I$ | 1/10/day | [24, 32] | $\eta_H$ | 0.25 (dimensionless) | Assumed |
| $\gamma_A$ | 1/5/day | [31] | $\xi_v$ | $2.97 \times 10^{-4}$/day | Assumed |
| $\gamma_H$ | 1/8/day | [24] | $\varepsilon_v$ | 0.70 (dimensionless) | [11, 12] |

**Figure 2:**
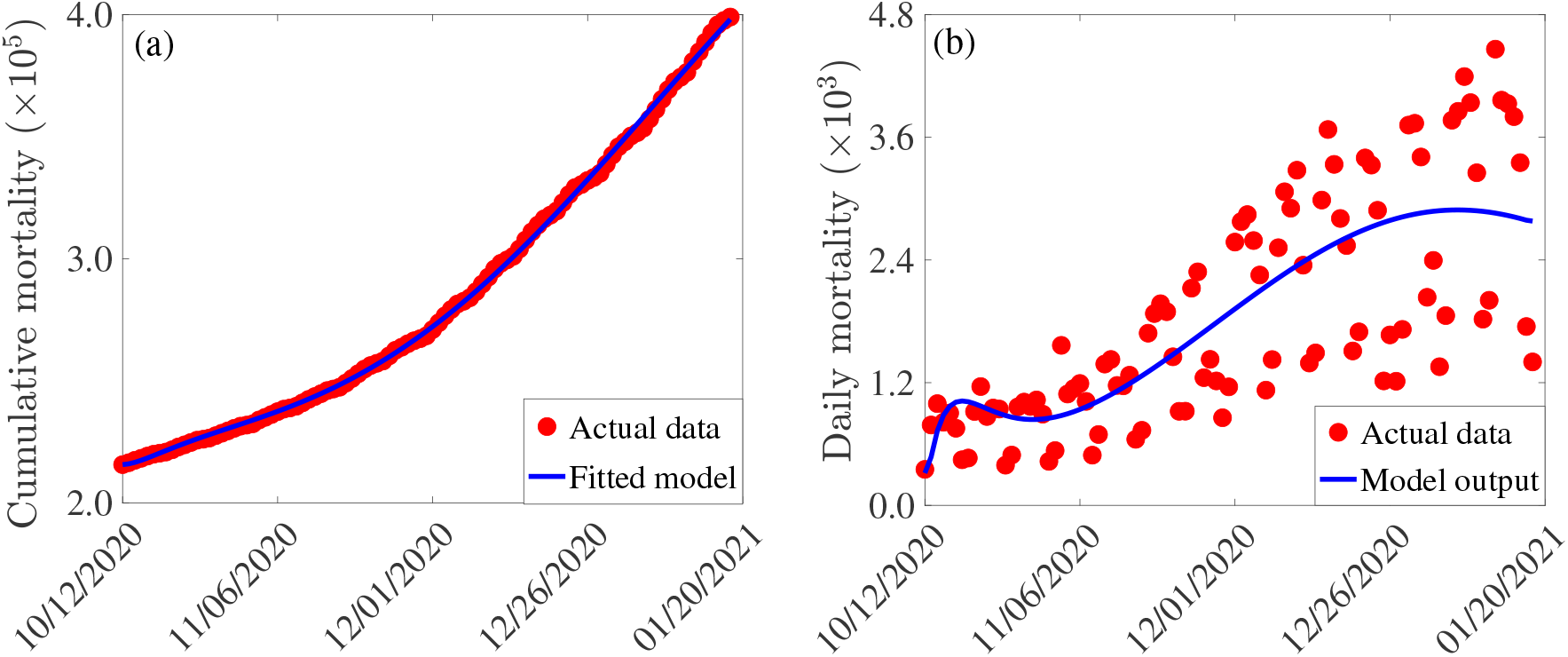
(a) Observed cumulative mortality (red dots), and the predicted cumulative mortality (blue curve) for the U.S. generated using the model (2.5)-(2.6) (in the absence of vaccination) for the period from October 12, 2020 to January 20, 2021. (b) Simulations of the model (2.5)-(2.6) (without vaccination) using the estimated (fitted) and fixed parameters tabulated in Tables 3 and 4, respectively.

## 3 Mathematical Analysis

Since the model {(2.5), (2.6)} monitors the temporal dynamics of human populations, all state variables and parameters of the model are non-negative. Consider the following biologically-feasible region for the model:

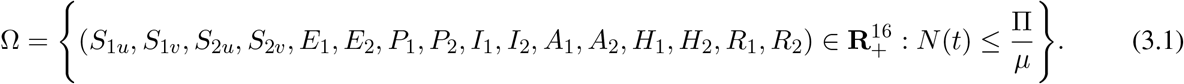

### Theorem 3.1.

*The region* Ω *is positively-invariant with respect to the model* {(2.5), (2.6)*}. Proof*. Adding all the equations of the model {(2.5), (2.6)*} gives*

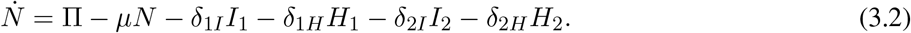

Recall that all parameters of the model {(2.5), (2.6)} are non-negative. Thus, it follows, from (3.2), that

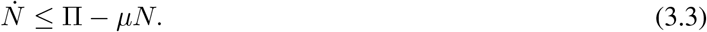

Hence, if 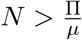, then 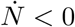. Furthermore, by applying a standard comparison theorem [33] on (3.3), we have:

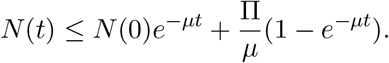

In particular, 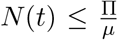 if 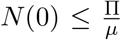. If 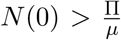 (i.e., *N* (0) is outside Ω), then 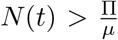, for all *t >* 0 but with 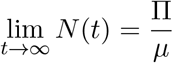 (and this type of solution trajectory strives to enter the region Ω). Thus, every solution of the model {(2.5), (2.6)} with initial conditions in Ω remains in Ω for all time *t >* 0. In other words, the region Ω is positively-invariant and attracts all initial solutions of the model {(2.5), (2.6)} . Hence, it is sufficient to consider the dynamics of the flow generated by {(2.5), (2.6)} in Ω (where the model is epidemiologically-and mathematically well-posed) [34].

### 3.1 Asymptotic Stability of Disease-free Equilibrium

The model {(2.5), (2.6)} has a unique disease-free equilibrium (DFE), obtained by setting all the infected compartments of the model to zero

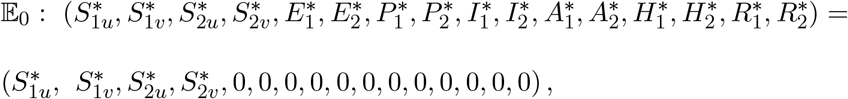

where,

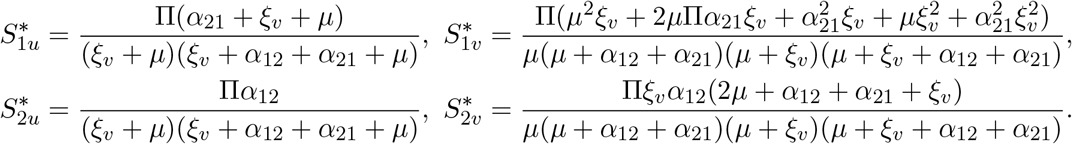

The local asymptotic stability property of the DFE (𝔼_0_) can be explored using the *next generation operator* method [35, 36]. In particular, using the notation in [35], it follows that the associated non-negative matrix (*F*) of new infection terms, and the M-matrix (*V*), of the linear transition terms in the infected compartments, are given, respectively, by (where the entries *f*_*i*_ and *g*_*i*_, *i* = 1, …, 8, of the non-negative matrix *F*, are given in Appendix I):

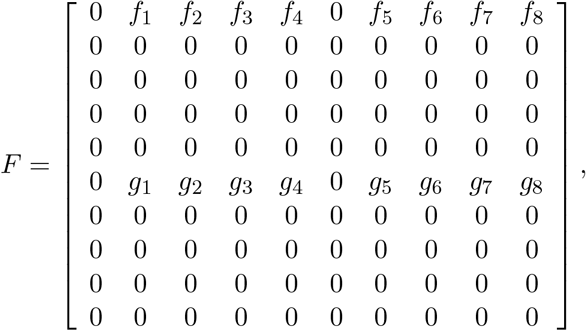

and,

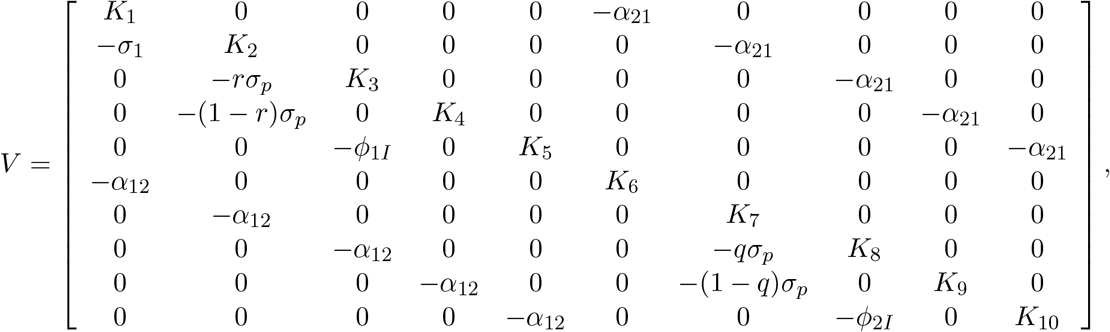

where *K*_1_ = *α*_12_ + *σ*_1_ + *µ, K*_2_ = *α*_12_ + *σ*_*P*_ + *µ, K*_3_ = *α*_12_ + *ϕ*_1*I*_ + *γ*_1*I*_ + *µ* + *δ*_1*I*_, *K*_4_ = *α*_12_ + *γ*_1*A*_ + *µ, K*_5_ = *α*_12_ + *γ*_1*H*_ + *µ* + *δ*_1*H*_, *K*_6_ = *α*_21_ + *σ*_2_ + *µ, K*_7_ = *α*_21_ + *σ*_*P*_ + *µ, K*_8_ = *α*_21_ + *ϕ*_2*I*_ + *γ*_2*I*_ + *µ* + *δ*_2*I*_, *K*_9_ = *α*_21_ + *γ*_2*A*_ + *µ* and *K*_10_ = *α*_21_ + *γ*_2*H*_ + *µ* + *δ*_2*H*_.

The theoretical analysis will be carried out for the special case of the model {(2.5), (2.6)} in the absence of the back-and-forth transitions between the no-mask and mask-user groups (i.e., the special case of the model with *α*_12_ = *α*_21_ = 0). This is needed for mathematical tractability. It follows that the *control reproduction number* of the model {(2.5), (2.6)} (with *α*_12_ = *α*_21_ = 0), denoted by ℛ_*c*_, is given by (where *ρ* is the spectral radius):

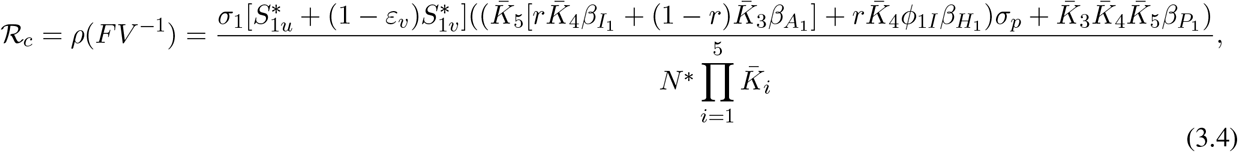

where, 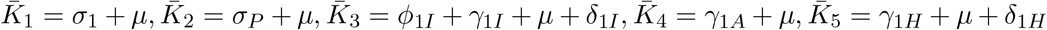. The result below follows from Theorem 2 of [35].

#### Theorem 3.2.

*The DFE (*𝔼_0_) *of the model* {(2.5), (2.6)}, *with α*_12_ = *α*_21_ = 0, *is locally-asymptotically stable if* ℛ_*c*_ *<* 1, *and unstable if* ℛ_*c*_ *>* 1.

The threshold quantity ℛ_*c*_ is the *control reproduction number* of the model {(2.5), (2.6)} . It measures the average number of new COVID-19 cases generated by a typical infectious individual introduced into a population where a certain fraction of the population is protected (*via* the use of interventions, such as face mask, social-distancing and/or vaccination). The epidemiological implication of Theorem 3.2 is that a small influx of COVID-19 cases will not generate an outbreak in the community if the control reproduction number (ℛ_*c*_) is brought to, and maintained at a, value less than unity. In the absence of public health interventions (i.e., in the absence of vaccination, face mask usage and social-distancing), the control reproduction number (ℛ_*c*_) reduces to the *basic reproduction number* (denoted by ℛ_0_), given by

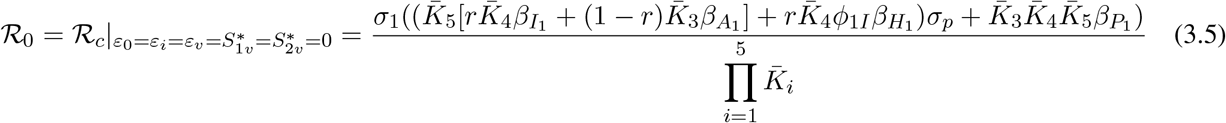

### 3.2 Derivation of Vaccine-induced Herd Immunity Threshold

Herd immunity is a measure of the minimum percentage of the number of individuals in a community that is susceptible to a disease that need to be protected (i.e., become immune) so that the disease can be eliminated from the population. There are two main ways to achieve herd immunity, namely through acquisition of natural immunity (following natural recovery from infection with the disease) or by vaccination. Vaccination is the safest and fastest way to achieve herd immunity [37, 38]. For vaccine-preventable diseases, such as COVID-19, not every susceptible member of the community can be vaccinated, for numerous reasons (such as individuals with certain underlying medical conditions, infants, pregnant women, or those who opt out of being vaccinated for various reasons etc.) [9]. So, the question, in the context of vaccine-preventable diseases, is what is the minimum proportion of individuals that can be vaccinated we need to vaccinate in order to achieve herd immunity (so that those individuals that cannot be vaccinated will become protected owing to the community-wide herd-immunity). In this section, a condition for achieving vaccine-derived herd immunity in the U.S. will be derived.

Let 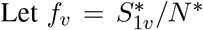, with *N* ^*∗*^ = Π*/µ*, be the proportion of susceptible individuals in Group 1 that have been vaccinated at the disease-free equilibrium (𝔼_0_). Using this definition in Equation (3.4) gives:

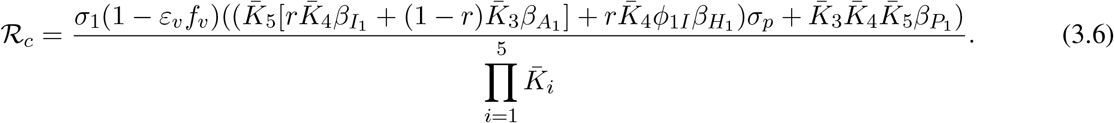

Setting ℛ_*c*_, in Equation (3.6), to unity and solving for *f*_*v*_ gives the herd immunity threshold (denoted by 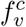) in terms of the basic reproduction number [9, 18]:

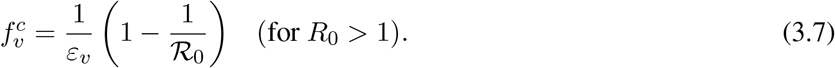

It follows from (3.6) and (3.7) that ℛ_*c*_ *<* (*>*)1 if 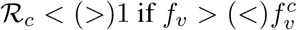. Further, ℛ_*c*_ = 1 whenever 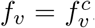. This result is summarized below:

#### Theorem 3.3.

*Consider the special case of the model* {(2.5), (2.6)} *with α*_12_ = *α*_21_ = 0. *Vaccine-induced herd immunity can be achieved in the U*.*S*., *using an imperfect anti-COVID vaccine, if* 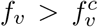 *(i*.*e*., *if* ℛ_*c*_ *<* 1*). If* 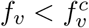 *(i*.*e*., *if* ℛ_*c*_ *>* 1), *then the vaccination program will fail to eliminate the COVID-19 pandemic in the U*.*S*.

The epidemiological implication of Theorem 3.3 is that the use of an imperfect anti-COVID vaccine can lead to the elimination of the COVID-19 pandemic in the U.S. if the sufficient number of individuals residing in the U.S. is vaccinated, such that 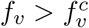. The Vaccination program will fail to eliminate the pandemic if the vaccine coverage level is below the aforementioned herd immunity threshold (i.e., if 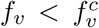). Although vaccination, no matter the coverage level, is always useful (i.e., vaccination will always reduce the associated reproduction number, ℛ_*c*_, thereby reducing disease burden, even if the program is unable to bring the reproduction number to a value less than unity), elimination can only be achieved if the herd immunity threshold is reached (i.e., disease elimination is only feasible if the associated reproduction number of the model is reduced to, and maintained at, a value less than unity). The pandemic will persist in the U.S. if ℛ_*c*_ *>* 1.

Figure 3(a) depicts the cumulative mortality of COVID-19 in the U.S. for various steady-state vaccination coverage levels (*f*_*v*_). This figure shows a decrease in cumulative mortality with increasing vaccination coverage. In particular, a marked decrease in cumulative mortality, in comparison to the baseline cumulative mortality (blue curve in Figure 3(a)), is recorded when herd immunity (i.e., when 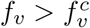) is attained (green curve of Figure 3(a)). While a noticeable decrease in the cumulative mortality is also observed when the vaccine coverage equals the herd immunity threshold (gold curve of Figure 3(a)), the cumulative mortality dramatically increases (in comparison to the baseline, depicted by the blue curve of this figure) if the vaccine coverage is below the herd immunity threshold (magenta curve of Figure 3(a)).

**Figure 3:**
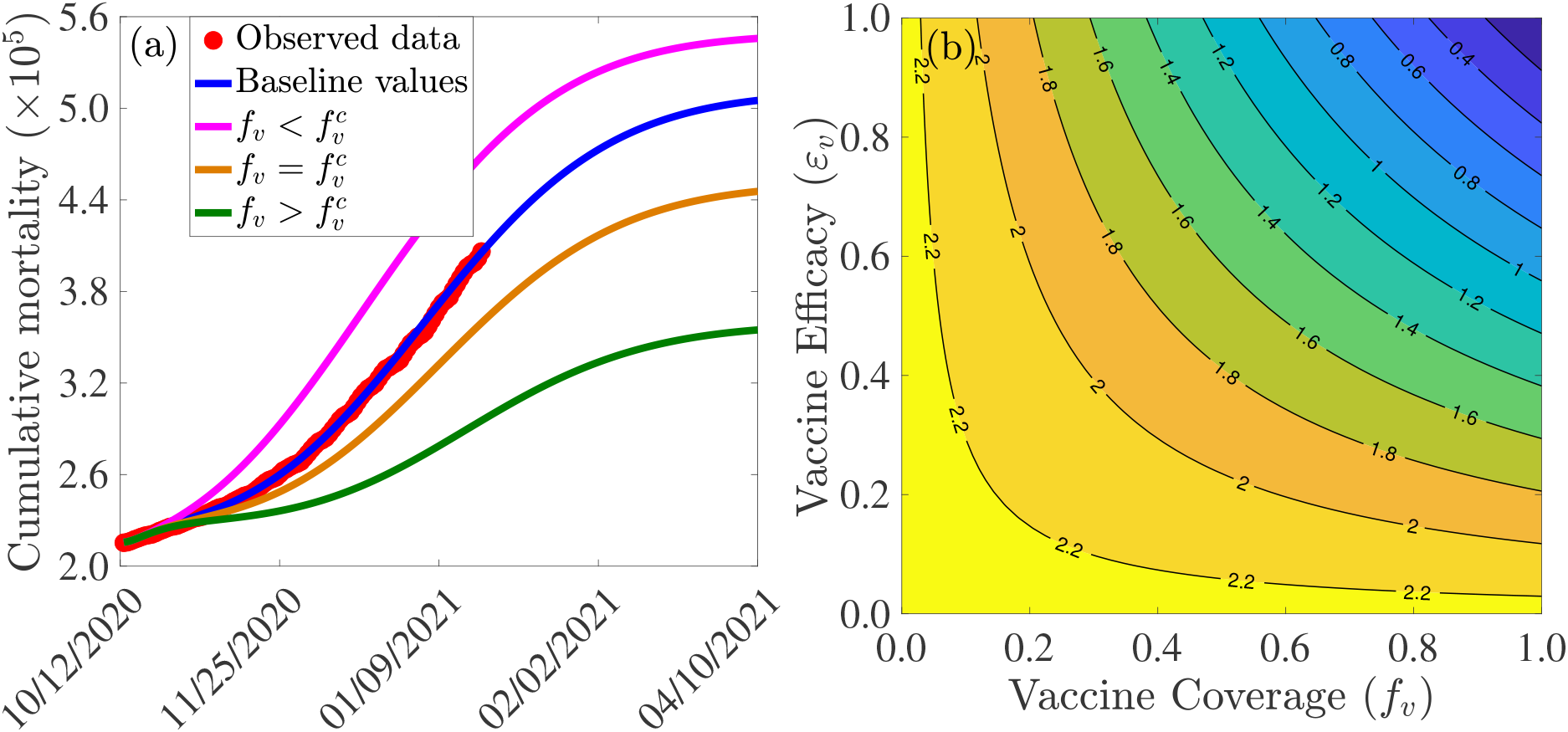
Assessment of the effects of vaccine coverage (*f*_*v*_) and efficacy (*ε*_*v*_) on COVID-19 dynamics in the U.S. (a) Simulations of the model {(2.5), (2.6)}, with *α*_12_ = *α*_21_ = 0, showing the cumulative COVID-19 mortality in the U.S., as a function of time, for various values of vaccine coverage. Parameter values used are as given by the baseline values in Tables 3-4, with *α*_12_ = *α*_21_ = 0 and various values of *f*_*v*_. Magenta curve 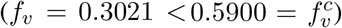, blue curve (baseline parameter values, and baseline level of social-distancing compliance inherent in the cumulative mortality data, for the period October 12, 2020 to January 20, 2021, used to fit the model), gold curve 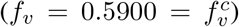 and green curve 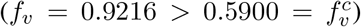. The observed cumulative deaths data, fitted to the baseline scenario predicted by the model (blue curve), is shown in red dots. (b) Contour plot of the control reproduction number (ℛ _*c*_) of the model {(2.5), (2.6)}, with *α*_12_ = *α*_21_ = 0, as a function of vaccine coverage (*f*_*v*_) and vaccine efficacy (*ε*_*v*_). Parameter values used are as given in Tables 3-4, with *α*_12_ = *α*_21_ = 0.

The effect of vaccination coverage (*f*_*v*_) and efficacy (*ε*_*v*_) on the control reproduction number (ℛ_*c*_) is assessed by depicted a contour plot of ℛ_*c*_, as a function of *f*_*v*_ and *ε*_*v*_. The results obtained (Figure 3(b)) shows that the values of the control reproduction number for the U.S., during the simulation period (October 12, 2020 to January 20, 2021), range from 0.4 to 2.2. Further, this figure shows that the control reproduction number decreases with increasing values of vaccination efficacy and coverage. For example, using the AstraZeneca vaccine (with efficacy *ε*_*v*_ = 0.7), about 80% of the U.S. population needs to be successfully vaccinated (with the two AstraZeneca doses) in order to bring the control reproduction number to a value less than unity. In other words, this figure shows that herd immunity can be achieved using the AstraZeneca vaccine in the U.S. if at least 80% of the populace received the two doses of the AstraZeneca vaccine. Using either the Pfizer or Moderna vaccine (each with efficacy of about 95%), on the other hand, the control reproduction number can be brought to a value less than unity (i.e., achieve herd immunity) if at least 60% of the U.S. populace received the two doses of either vaccine. Thus, this figure shows that the prospect of achieving vaccine-derived herd immunity using any of the three vaccines currently-available in the market (AstraZeneca, Pfizer and Moderna) is promising if the coverage is moderately-high enough (with the prospect far more likely to be achieved using the Pfizer or Moderna vaccine, in comparison to using the AstraZeneca vaccine).

We also explored the potential impact of additional social-distancing on the minimum vaccination coverage needed to achieve herd immunity. It should, first of all, be stressed that, since our model was parametrized using the cumulative mortality data during the third wave of the pandemic in the U.S. (October 12, 2020 to January 21, 2021), the effects of other nonpharmaceutical interventions, such as face masks usage and social-distancing, are already embedded into the results/data. In other words, the data (or the parametrization of our model) already includes some baseline level of these interventions. Specifically, we assume that the cumulative mortality data includes a baseline level of social-distancing compliance in the population (which is, clearly, quite high compared to what it was during the early stages of the pandemic in the U.S.) We now ask the question as to whether or not the minimum requirement for 80% and 60% coverage needed to achieve herd immunity, using the AstraZeneca or Pfizer/Moderna vaccine, respectively, can be reduced if the baseline social-distancing compliance is increased. In this study, we model social-distancing compliance by multiplying the effective contact rates (*β*_1_ and *β*_2_) with the factor 1 − *c*_*s*_, where 0 *< c*_*s*_ ≤1 is a measure of the additional social-distancing compliance (to the baseline social-distancing compliance achieved during the beginning of our simulation period; that is, by October 12, 2020).

We simulated the model {(2.5), (2.6)} using various values of *c*_*s*_, and the results obtained are tabulated in Table 5. This table shows that if an additional 5% of the U.S. population observe social-distancing in public (in addition to the baseline social-distancing compliance achieved by October 12, 2020), the minimum vaccine coverages required to achieve herd immunity using the AstraZeneca and Pfizer/Moderna vaccines reduce, respectively, to 77% and 56.4%. Furthermore, if the increase in baseline social-distancing compliance is 10%, the minimum coverage needed to achieve herd immunity further reduce (but marginally) to 73% and 54%, respectively. However, when the increase in baseline social-distancing compliance is 30%, herd immunity can be achieved using the AstraZeneca vaccine by vaccinating only 53% of the U.S. population with this vaccine. For this scenario, only about 39% of the U.S. population needs to be vaccinated to achieve herd immunity if either the Pfizer or Moderna vaccine is used.

**Table 5:** Vaccine-induced herd immunity threshold 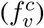 for the U.S. for various levels of increases in baseline social-distancing compliance (*c*_*s*_). Parameter values used are as given in Tables 3-4, with *α*_12_ = *α*_21_ = 0.

|  | Herd threshold | Herd threshold | Herd threshold | Herd threshold |
| --- | --- | --- | --- | --- |
| Vaccine name (efficacy) | $c_s = 0$ (baseline) | $c_s = 5\%$ | $c_s = 10\%$ | $c_s = 30\%$ |
| AstraZeneca ($\varepsilon_v = 70\%$) | $f_v^c = 80\%$ | $f_v^c = 77\%$ | $f_v^c = 73\%$ | $f_v^c = 53\%$ |
| Pfizer & Moderna ($\varepsilon_v = 95\%$) | $f_v^c = 59\%$ | $f_v^c = 56.4\%$ | $f_v^c = 54\%$ | $f_v^c = 39\%$ |

Thus, this study shows that the prospect of achieving vaccine-derived herd immunity in the U.S. using any of the three currently-available vaccines is greatly enhanced if the vaccination program is complemented with an increased (and sustained) social-distancing strategy (from the baseline). In other words, if more people living in the U.S. will continue to observe social-distancing (e.g., additional 30% from the baseline social-distancing compliance), then COVID-19 elimination can be achieved if roughly only half the population is vaccinated using the AstraZeneca vaccine, or 2 in 5 vaccinated if either the Pfizer or Moderna vaccine is used instead. The U.S. is currently using the latter vaccines. Hence, with about 30% additional social-distancing compliance, we would only need to vaccinate about 2 in 5 residents of the U.S. to achieve vaccine-derived herd immunity (hence, eliminate the pandemic).

## 4 Numerical Simulations: Assessment of Control Strategies

The model {(2.5), (2.6)} will now be simulated to assess the population-level impact of the various intervention strategies described in this study. In particular, our objective is to assess the impact of social-distancing and face mask usage, implemented as sole interventions and in combination with any of the three currently-available anti-COVID vaccines (namely the AstraZeneca, Moderna and Pfizer vaccines), on curtailing (or eliminating) the burden of the COVID-19 pandemic in the United States. Unless otherwise stated, the simulations will be carried out using the estimated (fitted) and fixed baseline values of the parameters of the model tabulated in Tables 3-4. Furthermore, unless otherwise stated, the baseline initial size of the population of individuals who habitually wear face masks in public (assumed to be 30%), denoted by *N*_2_(0), will be used in the simulations. The numerical simulation results for the baseline scenario (i.e., where baseline values of the parameters of the model, as well as the baseline initial size of the mask-wearing population, are used) will be illustrated in blue curves in the forthcoming figures. Furthermore, all numerical simulations will be carried out for the period starting from October 12, 2020 (which corresponds to the onset of the third wave of the pandemic in the United States).

### 4.1 Assessing the Impact of Initial Population of Face Mask Wearers

The model (2.5)-(2.6) is simulated to assess the community-wide impact of using face masks, as the sole intervention, in curtailing the spread of the pandemic in the United States. Specifically, we simulate the model using the baseline values of the parameters in Tables 3-4 and various values of the initial size of the population of individuals who habitually wear face masks in public since the beginning of the pandemic in the United States (denoted by *N*_2_(0)). It should be noted that the parameters associated with other interventions (e.g., vaccination-related and social-distancing-related parameters) are kept at their baseline values given in Tables 3-4. Although a sizable number of U.S. residents (notably individuals categorized in the first-tier priority group for receiving the COVID-19 vaccine, such as frontline healthcare workers, individuals at residential care facilities, the elderly etc.) have already been vaccinated using one of the two FDA-approved vaccines (20.54 million vaccines doses have already been administered in the U.S. as of January 23, 2021 [39]), these vaccines are not expected to be widely available to the general public until some time in March or April, 2021. Consequently, we set March 15, 2021 as our reference point for when we expect the vaccines to be widely available to the general public. Under this scenario (of vaccines expected to be widely available a few months after the initial starting point of our simulations, namely October 12, 2020), the objective of this set of simulations is to assess the impact of face masks usage, as a sole intervention, in controlling the spread of the pandemic in the U.S. before the two FDA-approved vaccines (Pfizer or Moderna) become widely available to the general U.S. public (to the extent that high vaccination coverage, such as vaccinating one million U.S. residents *per* day, can be realistically achieved). The new U.S. administration aims to vaccinate 100 million residents during its first 100 days.

The simulation results obtained, depicted in Figure 4, show (generally) that the early adoption of face masks control measures (as measured in terms of the initial proportion of the populace who choose to habitually wear face masks whenever they are out in the public, denoted by *N*_2_(0)) play a vital role in curtailing the COVID-19 mortality in the U.S., particularly for the case when mask-wearers do not abandon their masks-wearing habit (i.e., *α*_21_ = 0). For the case where the parameters associated with the back-and-forth transitions between the masking and non-masking sub-populations (i.e., *α*_12_ and *α*_21_) are maintained at their baseline values (given in Tables 3-4), this figure shows that the size of the initial proportion of individuals who wear face masks has a significant impact on the cumulative COVID-19 mortality, as measured in relation to the cumulative mortality recorded when the initial proportion of mask wearers is at baseline level (blue curves in Figure 4). In particular, a 34% reduction in the cumulative mortality, in comparison to the cumulative mortality for the baseline scenario, will be recorded by March 15, 2021, if the initial proportion of mask-wearers is 40% (Figure 4(a), magenta curve). Furthermore, the reduction in cumulative mortality by March 15, 2021 increases to 52% if the initial proportion of mask-wearers is 75% (Figure 4(a), green curve). On the other hand, for the case when mask-wearers remain mask-wearers since the beginning of the simulation period (i.e., since October 12, 2020), so that *α*_21_ = 0, while non-mask wearers (i.e., those in Group 1) can change their behavior and become mask-wearers (i.e., *α*_12_ ≠ 0), our simulations show that the initial proportion of individuals who adopt masking only marginally affects the cumulative mortality (Figure 4(b)), in relation to the scenario in Figure 4(a), where both *α*_12_ and *α*_21_ are nonzero). In particular, if 40% of the U.S. population adopted mask-wearing right from the aforementioned October 12, 2020, up to 37% of the baseline COVID-19 mortality can be averted (Figure 4(b), magenta curve), in comparison to the baseline (Figure 4(b), blue curve). Furthermore, the reduction in baseline cumulative mortality rises to 53% if three in every four Americans opted to wear face masks since the beginning of the simulation period (Figure 4(b), green curve). This also represents a marginal increase in the cumulative deaths averted, in comparison to the scenario when *α*_12_ ≠ 0 and *α*_21_ ≠ 0 (Figure 4(a), green curve).

**Figure 4:**
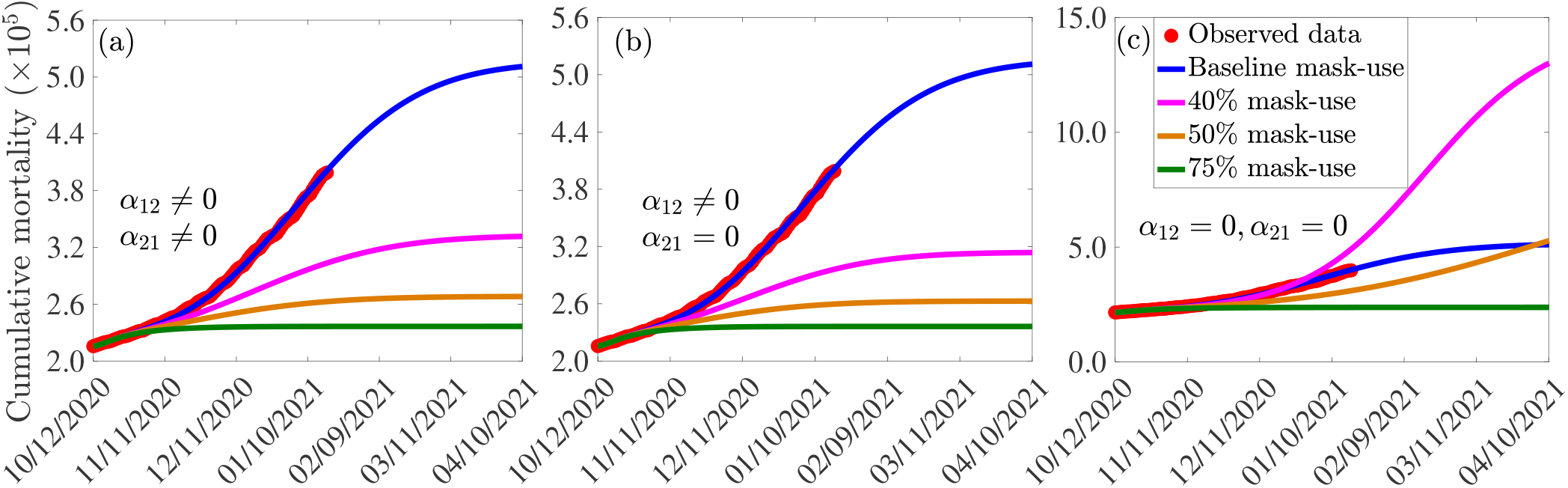
Assessment of the impact of face mask usage, as a sole intervention, on COVID-19 pandemic in the U.S. Simulations of the model (2.5)-(2.6), showing cumulative mortality, as a function of time, for (a) face mask transition parameters (*α*_12_ and *α*_21_) maintained at their baseline values, (b) mask-wearers strictly adhere to wearing masks (*α*_21_ = 0) and non-mask-wearers transit to mask wearing at their baseline rate (*α*_12_ ≠ 0), and (c) non-mask wearers and mask-wearers do not change their behavior (i.e., *α*_12_ = *α*_21_ = 0). Mask use change is implemented in terms of changes in the initial size of the population of individuals who wear face masks (from the onset of simulations, on October 12, 2020). Blue curves (in each of the plots) represent the baseline scenarios where the initial size of the population of mask wearers is fixed at 30%, and the transition parameters, *α*_12_ and *α*_21_, are maintained at their baseline values. Parameter values used in the simulations are as given by the baseline values in Tables 3-4, with different values of *α*_12_ and *α*_21_ (except for the blue curves, where *α*_12_ and *α*_21_ are fixed at their baseline values).

For the case when no back-and-forth transitions between the two (mask-wearing and non-mask-wearing) groups is allowed (i.e., when *α*_12_ = *α*_21_ = 0), our simulations show a far more dramatic effect of face mask usage on COVID-19 mortality (Figure 4(c)). For instance, this figure shows that higher cumulative mortality is recorded, in comparison to the baseline masks use scenario, when the initial size of the population of mask wearers is 40% (Figure 4(c), magenta curve), in comparison to the blue curve of the same figure). Specifically, this represents a 55% increase, in comparison to the baseline cumulative mortality. This simulation result suggests that the 40% initial size of the populace wearing face masks, during the onset of the third wave of the pandemic in the U.S. (starting October 12, 2020), falls below the mask-use compliance threshold level needed to reduce the cumulative mortality during the third wave. On the other hand, if the initial size of the population of face masks wearers is increased to 50%, a decrease (and not an increase) in cumulative mortality is recorded, in comparison to the cumulative mortality for the baseline scenario (Figure 4(c),gold curve, in comparison to the blue curve of the same figure). Further dramatic reduction (52%), in relation to the baseline scenario, will be achieved if the initial size of the mask-wearing population is increased to 75% (Figure 4(c), green curve, in comparison to the blue curve of the same figure). Thus, these simulations show that, for the case when no change of mask-wearing behavior is allowed (i.e., everyone remains in their original group), there is a threshold value of the initial size of the population of mask wearers above (below) which the cumulative mortality is decreased (increased). Specifically, this simulation shows that (for this scenario with *α*_12_ = *α*_21_ = 0), at least half the population need to be wearing face masks right from the beginning of the epidemic to ensure greater reduction in cumulative mortality, in comparison to the baseline scenario (when the initial size of the mask-wearing sub-population is 30%).

In summary, comparing the same initial mask coverage (i.e., the same curve colors) in Figures 4(a)-(c), it is clear that the scenario where individuals are allowed to change their behaviors from not wearing face masks to wearing face masks (i.e., *α*_12_ ≠ 0), but masks wearers do not abandon masks wearing (i.e., *α*_21_ = 0), depicted in Figure 4(b), resulted in saving more lives (*albeit* only slightly), compared to the scenarios where no change of behavior is allowed for members of each group (Figure 4(c)) or members of both groups can change their behavior (Figure 4(a)). In other words, out study emphasize the need for non-maskers to adopt a mask-wearing culture (i.e., *α*_12_ = 0) and habitually masks wearers do not abandon their mask-wearing habit (i.e., *α*_21_ = 0).

### 4.2 Assessing the Impact of Additional Social-distancing Compliance

In this section, we carry out numerical simulations to assess the potential impact of increases in the baseline social-distancing compliance (*c*_*s*_) on the control of the pandemic. Specifically, the model {(2.5), (2.6)} will be simulated using the baseline parameter values tabulated in Tables 3-4 with various values of *c*_*s*_ (corresponding to the various levels of the increase in baseline social-distancing compliance in the U.S., starting from October 12, 2020). It should be noted that, for these simulations, the baseline initial size of the masking population, *N*_2_(0), is maintained. Furthermore, vaccine-related parameter values are maintained at their baseline levels in Tables 3-4.

The simulation results obtained, depicted in Figure 5, show that, in the absence of additional increase in baseline social-distancing (i.e., *c*_*s*_ = 0, so that social-distancing compliance is maintained at the baseline level inherent in the cumulative mortality data by October 12, 2020), the U.S. would record about 500,000 cumulative deaths by March 15, 2021 (Figure 5(a), blue curve). For this (baseline social-distancing) scenario, the U.S. would have recorded a peak daily mortality of about 3, 000 deaths on January 5, 2021 (Figure 5(b), blue curve). The simulations in Figure 5 further show that the cumulative mortality (Figure 5(a)) and daily mortality (Figure 5(b)) decrease with increasing levels of the additional social-distancing compliance (*c*_*s*_) in the population. For example, if the baseline social-distancing achieved during the onset of the third wave of the pandemic in the U.S. is further increased by only 5%, the simulation results show that up to a 19% of the cumulative mortality can be averted by March 15, 2021 (Figure 5(a), magenta curve), in comparison to the baseline social-distancing scenario (Figure 5(a), blue curve). Similarly, for this 5% increase in social-distancing (in relation to the baseline), up to 36% reduction in daily mortality can be achieved (Figure 5(b), magenta curve), in comparison to the baseline scenario (Figure 5(b), blue curve), and the pandemic would have peaked a week earlier (in late December 2020; the daily mortality at this peak would have been 1, 900), in comparison to the peak recorded in the baseline social-distancing scenario ((Figure 5(b), blue curve). More dramatic reduction in mortality will be recorded if the level of additional socialdistancing compliance is further increased. For instance, if the baseline social-distancing compliance is increased by 10%, our simulations show that about 31% of the cumulative deaths recorded for the case with baseline socialdistancing scenario ((Figure 5(a), blue curve) would have been averted (Figure 5(a), gold curve). For this scenario, up to 59% of the daily deaths would have been prevented and the pandemic would have peaked in mid December 2020 (the daily mortality at this peak would have been 1, 229), as depicted in the gold curve of Figure 5(b). Finally, if the baseline social-distancing compliance is increased by 30%, the pandemic would have failed to generate a major outbreak in the U.S. (Figure 5, green curves). In particular, the cumulative mortality for the U.S. by March 15, 2021 will be about 252, 400 (as against the nearly 400,000 fatalities that were recorded), as shown by the green curve of Figure 5(a), in comparison to the blue curve of the same figure.

**Figure 5:**
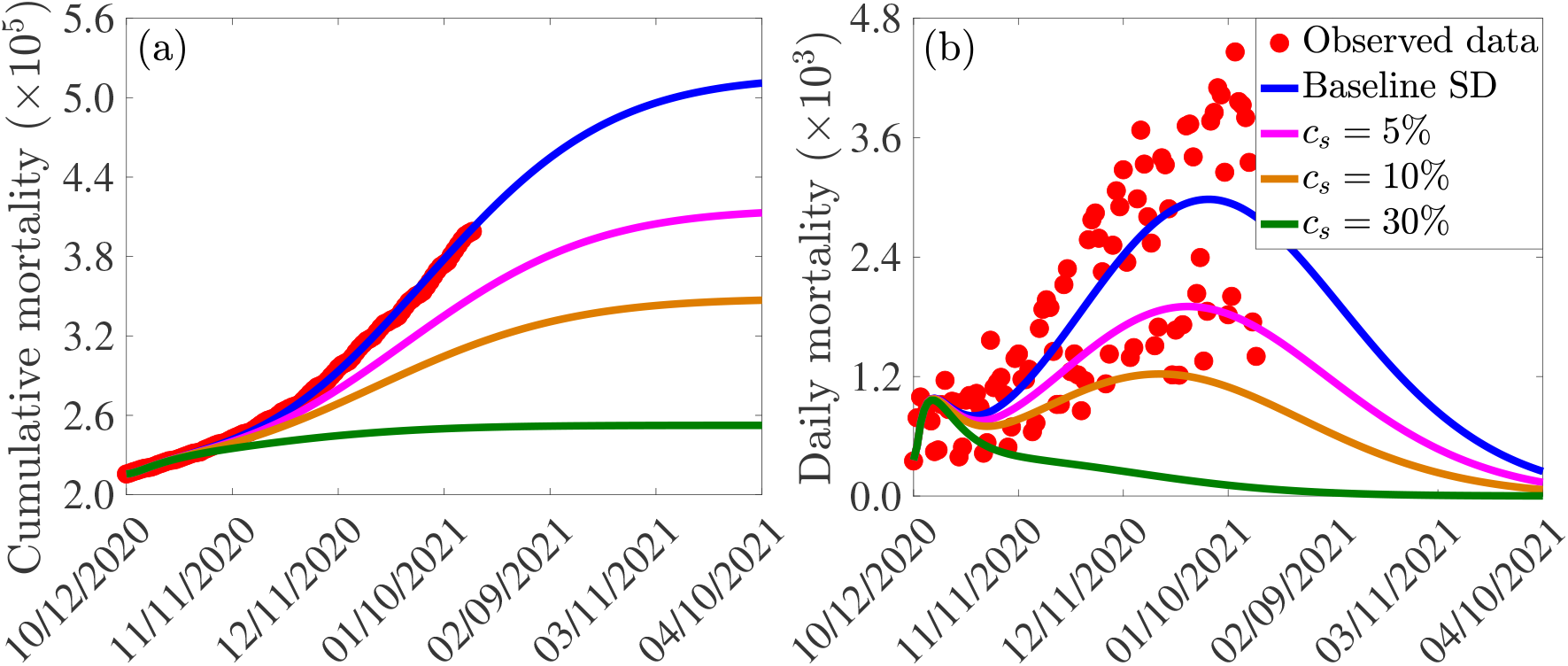
Assessment of the singular impact of increases in baseline social-distancing compliance on COVID-19 pandemic in the U.S. Simulations of the model (2.5)-(2.6) showing (a) cumulative mortality, as a function of time; (b) daily mortality, as a function of time, for various levels of increases in baseline social-distancing (SD) compliance (*c*_*s*_) attained during the third wave of the pandemic in the United States. Parameter values used in the simulations are as given by the baseline values in Tables 3-4, with *β*_1_ and *β*_2_ multiplied by (1 − *c*_*s*_).

In summary, the results in Figure 5 show that COVID-19 could have been effectively suppressed in the U.S. if the baseline social-distancing compliance (recorded during the onset of the third wave of the pandemic in early October 2020) is increased by about 10% to 30%. These (recommended) increases in social-distancing compliance seem reasonably attainable. Hence, our study suggests that a moderate increase in the baseline social-distancing compliance will lead to the effective control of the COVID-19 pandemic in the U.S. This (increase in baseline social-distancing, as well as face masks usage) should be sustained until herd immunity is attained.

### 4.3 Assessment of Combined Impact of Vaccination and Social-distancing

The model (2.5)-(2.6) will now be simulated to assess the community-wide impact of the combined vaccination and social-distancing strategy. Although the two FDA-approved vaccines were approved for use by mid December 2020, we assume a hypothetical situation in which the vaccination started by mid October 2020 (the reason is to ensure consistency with the cumulative mortality data we used, which started from October 12, 2020 corresponding to the onset of the third wave of the pandemic in the United States). We consider the three vaccines currently being used in humans, namely the AstraZeneca vaccine (with estimated efficacy of 70%) and the two FDA-approved vaccines (Moderna and Pfizer vaccines, each with estimated efficacy of about 95%). Simulations are carried out using the baseline parameter values in Tables 3-4, with various values of the vaccination coverage parameter (*ξ*_*v*_). For these simulations, parameters and initial conditions related to the other intervention (face mask usage) are maintained at their baseline values. Since the Moderna and Pfizer vaccines have essentially the same estimated efficacy (≈95%), we group them together in the numerical simulations for this section.

The simulation results obtained for the Moderna and Pfizer vaccine, depicted in Figures 6(a)-(c)), show that, in the absence of vaccination (and with social-distancing at baseline compliance level), approximately 511, 100 cumulative deaths will be recorded in the U.S. by April 10, 2021 (blue curves of Figures 6(a)-(c)). Furthermore, this figure shows a marked reduction in daily mortality with increasing vaccination coverage (*ξ*_*v*_). This reduction further increases if vaccination is combined with social-distancing. For instance, with social-distancing compliance maintained at its baseline value on October 12, 2020 (i.e., *c*_*s*_ = 0), vaccinating at a rate of 0.00074 *per* day (which roughly translates to vaccinating 250, 000 people every day) resulted in a reduction of the projected cumulative mortality recorded by April 10, 2021 by 12%, in comparison to the case when no vaccination is used (magenta curve in Figure 6(a), in comparison to the blue curve of the same figure). In fact, up to 31% of the projected cumulative mortality to be recorded by April 10, 2021 could be averted if, for this vaccination rate, the baseline social-distancing compliance is increased by 10% (i.e., *c*_*s*_ = 0.1; magenta curve in Figure 6(c), in comparison to magenta curve in Figure 6(a)). If the vaccination rate is further increased to, for instance, *ξ*_*v*_ = 0.0015 *per* day (corresponding to vaccinating about 500, 000 people every day), while keeping social-distancing at its baseline compliance level (i.e., *c*_*s*_ = 0), our simulations show a reduction of 27% in the projected cumulative mortality by April 10, 2021, in comparison to the baseline social-distancing scenario (gold curve, Figure 6(a), in comparison to the blue curve of the same figure). This reduction increases to 38% if the vaccination program is supplemented with social-distancing that increases the baseline compliance by 10% (gold curve, Figures 6(c)). If 1 million people are vaccinated *per* day (i.e., *ξ*_*v*_ = 0.003 *per* day), our simulations show that the use of the Moderna and Pfizer vaccines could lead to up to 36% reduction in the projected cumulative mortality by April 10, 2021 in the U.S. if the vaccination program is combined with a 10% increase in social-distancing compliance level (green curve of Figure 6(c)). Finally, compared to the Moderna and Pfizer vaccines, slightly lower reductions in the projected cumulative mortality are recorded when the AstraZeneca vaccine (with moderate to high vaccination coverage) is used (Figures 6(d)-(f)), particularly if combined with social-distancing. These results are summarized in Table 6.

**Figure 6:**
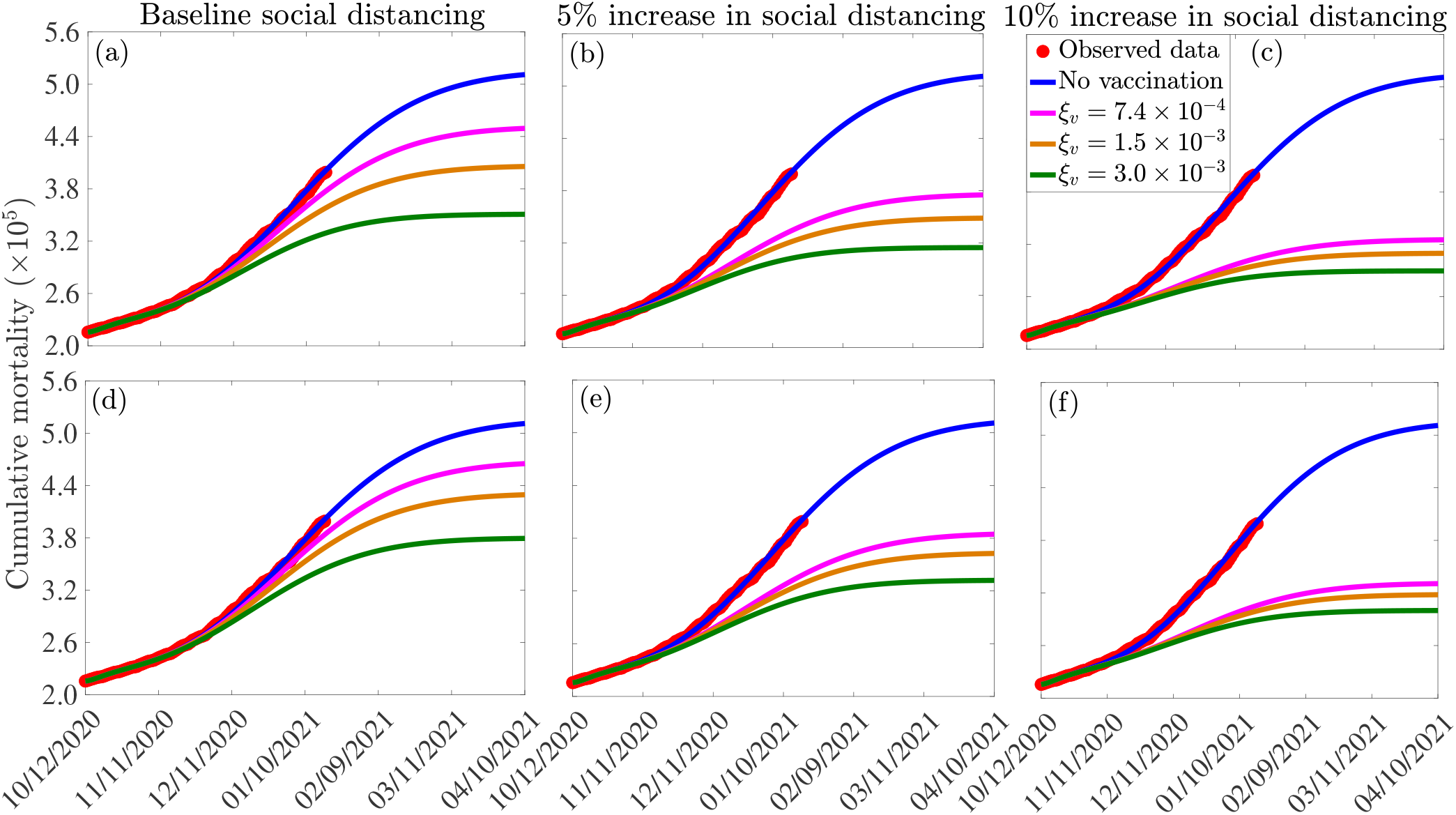
Assessment of the combined impact of vaccination and social-distancing on cumulative mortality. Simulations of the model (2.5)-(2.6), depicting cumulative as a function of time, for the currently available anti-COVID-19 vaccines and various levels of increases in baseline social-distancing compliance starting from October 12, 2020 (*c*_*s*_). (a)-(c): Pfizer or Moderna vaccine. (d)-(f): AstraZeneca vaccine. The vaccination rates *ξ*_*v*_ = 7.4 × 10^*−*4^, 1.5 × 10^*−*3^ *per* day, and 3.0 × 10^*−*3^ *per* day correspond, respectively, to vaccinating approximately 2.5 × 10^5^, 5.0 × 10^5^ and 1.0 × 10^6^ people *per* day. Other parameter values of the model used are as presented in Tables 3-4.

**Table 6:** Percentage reduction in projected cumulative COVID-19 mortality on April 10, 2021, in relation to the cumulative mortality in the absence of vaccination (511, 100 COVID-19 deaths on April 10, 2021), for the three currently-available anti-COVID-19 vaccines: AstraZeneca vaccine (efficacy *ε*_*v*_ = 0.7); Pfizer and/or Moderna vaccine (efficacy *ε*_*v*_ = 0.95), and various levels of increases in baseline social-distancing compliance attained on October 12, 2020 (*c*_*s*_) and vaccination rate (*ξ*_*v*_). Notation: SD represents social-distancing compliance.

| Number of people vaccinated *per day* | Reduction with<br>Baseline SD ($c_s = 0$) | | Reduction with<br>$c_s = 0.05$ | | Reduction with<br>$c_s = 0.10$ | |
| --- | --- | --- | --- | --- | --- | --- |
| | $\varepsilon_v = 70\%$ | $\varepsilon_v = 95\%$ | $\varepsilon_v = 70\%$ | $\varepsilon_v = 95\%$ | $\varepsilon_v = 70\%$ | $\varepsilon_v = 95\%$ |
| 250,000 | 9% | 12% | 25% | 27% | 35% | 36% |
| 500,000 | 16% | 20% | 29% | 32% | 38% | 39% |
| 1,000,000 | 26% | 31% | 35% | 38% | 41% | 43% |

### 4.4 Combined Impact of Vaccination and Social-distancing on Time-to-elimination

The model (2.5)-(2.6) will now be simulated to assess the population-level impact of the combined vaccination and social-distancing interventions on the expected time the pandemic might be eliminated in the U.S. if the two strategies are implemented together. Mathematically, we define “elimination” to mean when the number of daily new cases is identically zero. As in Section 4.3, we consider the three currently-available vaccines (AstraZeneca, Moderna and the Pfizer vaccines), and assume that the vaccination program was started on October 12, 2020. The model is simulated to generate a time series of new daily COVID-19 cases in the U.S., for various vaccination rate (*ξ*_*v*_) and levels of increases in baseline social-distancing compliance (*c*_*s*_).

The results obtained, for each of the three currently-available vaccines, are depicted in Figures 7. This figure shows a marked decrease in disease burden (measured in terms of the number of new daily cases), with the possibility of elimination of the pandemic within 8 − 10 months from the commencement of the vaccination program. In particular, these simulations show that vaccinating 250, 000 people *per* day, with the Moderna or the Pfizer vaccine, will result in COVID-19 elimination in the U.S. by mid August of 2021, if the social-distancing compliance is kept at its current baseline compliance level (blue curve of Figure 7(a)). For this scenario, the elimination will be reached in late August 2021 using the AstraZeneca vaccine. If the vaccination rate is further increased, such as to vaccinating 1 million people every day (and keeping social-distancing at its October 12, 2020 baseline), COVID-19 elimination is achieved much sooner in the United States. For instance, for this scenario (i.e., with *ξ*_*v*_ = 0.003 *per* day), the pandemic can be eliminated by late June of 2021 using the Moderna or the Pfizer vaccines (green curve of Figure 7(a)) and by mid July of 2021 using the AstraZeneca vaccine (blue curve of Figure 7(d)).

**Figure 7:**
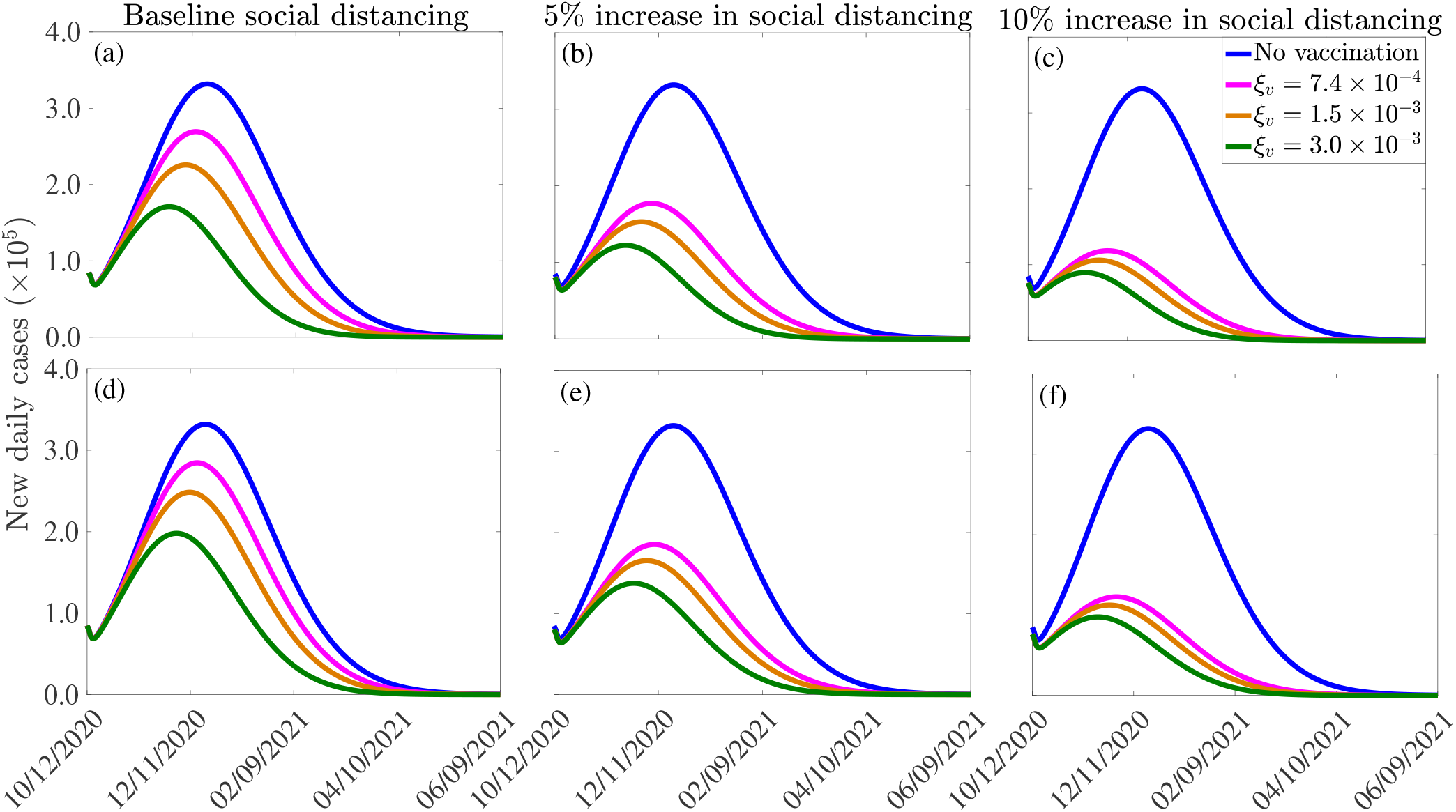
Effect of vaccination and social-distancing on time-to-elimination. Simulations of the model (2.5)-(2.6), depicting the impact of three currently-available vaccines against COVID-19 (the AstraZeneca vaccine, and the Pfizer or Moderna vaccine) and social-distancing, on time-to-elimination of the pandemic in the U.S. (a)-(c): Moderna or Pfizer vaccines. (d)-(f): AstraZeneca vaccine. The social-distancing compliance is baseline for (a) and (d), *c*_*s*_ = 0.05 for (b) and (e), and *c*_*s*_ = 0.10 for (e) and (f). The vaccination rates *ξ*_*v*_ = 7.4 × 10^*−*4^, 1.5 × 10^*−*3^, 3.0 × 10^*−*3^ *per* day correspond, respectively, to vaccinating approximately 2.5 × 10^5^, 5.0 × 10^5^, 1.0 × 10^6^ people *per* day. The values of the other parameters of the model used in the simulation are as given in Tables 3-4.

Our simulations further show that if the vaccination program is combined with social-distancing that increases the baseline compliance by 10%, COVID-19 can be eliminated in the U.S. by as early as the end of May of 2021 using the Moderna or the Pfizer vaccine (green curve of Figure 7(c)), and by late June of 2021 using the AstraZeneca vaccine (green curve, Figure 7(f)). In conclusion, these simulations show that any of the three currently-available vaccines considered in this study will lead to the elimination of the pandemic in the U.S. if the vaccination rate is moderately-high enough. The time-to-elimination depends on the vaccination rate and the level of increases in the baseline social-distancing compliance attained by October 12, 2020. The pandemic can be eliminated as early as the end of May of 2021 if moderate to high vaccination rate (e.g., 1 million people are vaccinated *per* day) and social-distancing compliance (e.g., *c*_*s*_ = 0.1) is attained and maintained.

It is worth mentioning that the two vaccines that are currently in used in the U.S. were only approved by the FDA in December 2020 (the Pfizer vaccine was approved on December 11, 2020, while the Moderna vaccine was approved a week later), and their administration into the arms of Americans started late in December 2020. Therefore, as we noted earlier, the greater U.S. community might only be able to receive any of the vaccines by March or April 2021 (we chose March 15, 2021 as our reference point for simulation/comparative purposes). Thus, with a mass vaccination start date of mid March 2021 (i.e., if we can only achieve vaccinating 1 million or more people daily from mid March 2021), then COVID-19 elimination, assuming a 10% increase in baseline social-distancing compliance achieved on October 12, 2020, can be achieved by the end of October 2021 using the Moderna or the Pfizer vaccine (for the AstraZeneca vaccine, elimination will extend to November of 2021). It should be mentioned that the elimination can be achieved even earlier if large scale community vaccination in the U.S. is started earlier than our projected March 15, 2021, and particularly if this (early large scale vaccination before March 15, 2021) is also complemented with significant increase in baseline social-distancing compliance (such as increasing the baseline compliance by 10%).

In summary, our study clearly shows that the prospect of eliminating COVID-19 in the U.S. by the middle or early fall of 2021 is very much feasible if moderate level of coverage can be achieved using either of the two vaccines being used in the U.S., and if this vaccination coverage is complemented with a social-distancing strategy that increases the baseline compliance achieved by October 12, 2020 by a mere 10%. Our study certainly points to the fact that we will be seeing the back of this devastating Coronavirus beast, and socio-economic life may return to near normalcy, in 2021.

### 4.5 Assessing the impacts of waning immunity, mask fatigue and relaxation of mask mandate for fully-vaccinated individuals, and therapeutic benefits of vaccines

In this section, the multi-group model (2.5)-(2.6) will be adapted and simulated to assess the population-level impact of three other factors that may significantly affect the effectiveness of the vaccination program against COVID-19, namely (a) waning natural and vaccine-derived immunity [40–42], (b) mask fatigue (and giving up masking) by fully-vaccinated individuals [43] and (c) therapeutic benefits of the vaccines (such as reducing development of severe disease, hospitalization and mortality in breakthrough infections, as well as in reducing transmissibility of infected vaccinated individuals) [15, 16, 44]. Although the model (2.5)-(2.6) does not explicitly incorporate the aforementioned factors, it can readily be adapted to allow for their assessment. We describe below how the model can be adapted to achieve this objective, in addition to illustrating the effects of the factors *via* numerical simulations of the resulting adapted version of the model (2.5)-(2.6). For consistency, the simulations in this section will also be carried out from the beginning of the third pandemic wave in the US (i.e., from October 12, 2020).

#### 4.5.1 Waning natural and vaccine-derived immunity

Waning natural immunity can be incorporated in the model by allowing a transition from the compartment of recovered individuals (for each of the two groups) into the corresponding compartment for unvaccinated susceptible individuals (i.e., the immunity derived from natural recovery from COVID-19 infection ultimately wanes, and the recovered individuals subsequently become wholly-susceptible again). To adapt the model to account for this, we introduce a new parameter, *ω*_*r*_, to represent the *per capita* rate at which recovered individuals revert to the corresponding unvaccinated susceptible compartment (i.e., the quantity *ω*_*r*_ *R*_*i*_, with *i* = 1, 2) is subtracted from the equation for *R*_*i*_ and added to the corresponding equation for *S*_*iu*_ in the model (2.5)-(2.6)).

Similarly, vaccine-derived waning immunity can be incorporated into the model (2.5)-(2.6) by allowing for transitions from the vaccinated susceptible compartments (*S*_*iv*_; *i* = 1, 2) to the corresponding unvaccinated susceptible compartment (*S*_*iu*_; *i* = 1, 2). We introduce a new parameter, *ω*_*v*_, to represent the rate of waning of vaccine-derived immunity. To incorporate this into the model, the quantity *ω*_*v*_*S*_*iv*_ (*i* = 1, 2) is subtracted from the equation for *S*_*iv*_ and added to the corresponding equation for *S*_*iu*_ in the model (2.5)-(2.6). For simulation purposes, we set *ω*_*r*_ and *ω*_*v*_ to be 1*/*270 *per* day and 1*/*180 *per* day [40, 42], respectively (corresponding to a nine and six months duration for the waning of natural and vaccine-derived immunity, respectively).

The model (2.5)-(2.6) is now simulated, using the parameter values in Tables 3 and 4, together with the above modifications (accounting for waning natural and vaccine-derived immunity, using the estimated values of *ω*_*r*_ and *ω*_*v*_), to assess the potential impact of waning immunity on the COVID-19 dynamics in the US. The results obtained, depicted in Figure 8(a), show a slight increase in the peak number of new daily cases, in comparison to the results in Figure 7(a), where the effect of waning immunity was not considered. In particular, if the vaccination rate is 250, 000 *per* day (i.e., if *ξ*_*v*_ is set at *ξ*_*v*_ = 7.4 × 10^*−*4^ *per* day), then the peak number of new cases increases by approximately 2% (in comparison to the case where no waning immunity is considered), and the time-to-elimination of the pandemic increases by about 13 days (compare blue curves in Figures 7(a) and 8(a)). If the daily vaccination rate is increased to one million *per* day (i.e., if *ξ*_*v*_ = 3.0 × 10^*−*3^ *per* day), then the peak new cases increases by up to 6% (in comparison to the case with no waning immunity) and the time-to-elimination increases by about a month (compare green curves in Figures 7(a) and 8(a)). The increases in burden and time-to-extinction in this case (with 1 million vaccinated daily, in comparison to the case with 250,000 people getting vaccinated daily) is due to the fact waning of both natural and vaccine-derived immunity causes a corresponding increase in the pool of susceptible individuals who can acquire infection (thereby increasing number of new cases and extending time-to-elimination). Thus, these simulations show that waning of natural and vaccine-derived immunity cause only a marginal increase in the burden and time-to-elimination of the pandemic.

**Figure 8:**
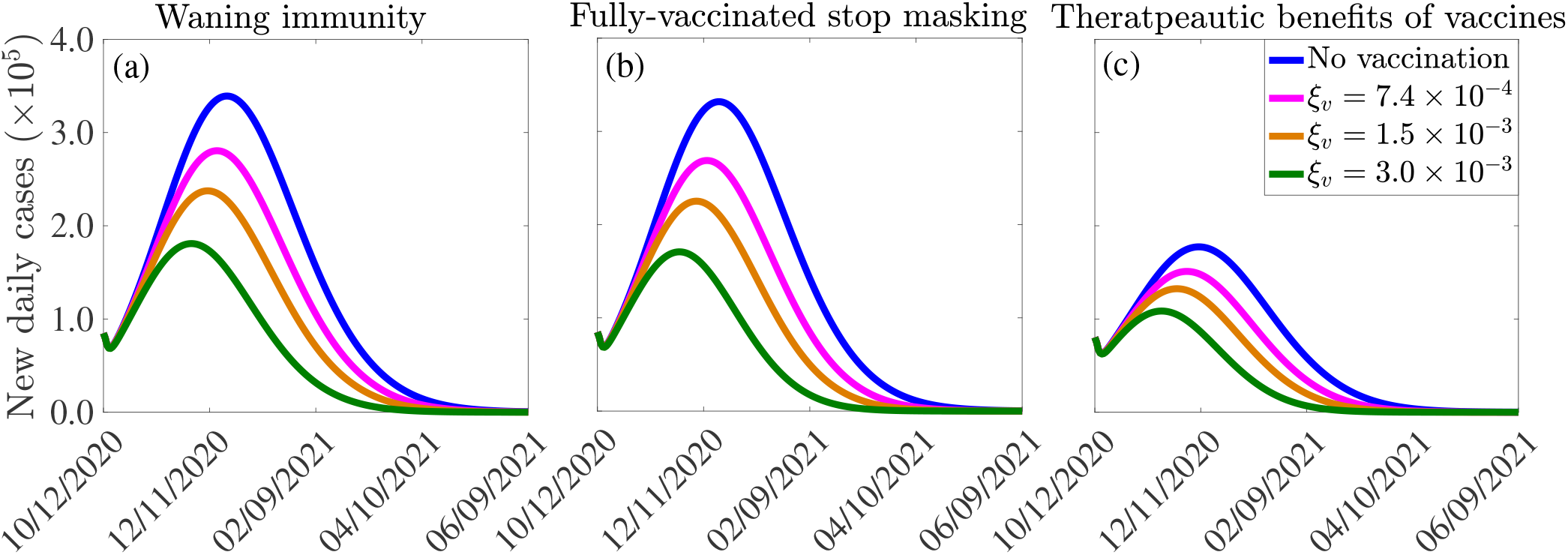
Effect of (a) vaccine-induced and natural immunity waning, (b) unmasking by vaccinated individuals, and (c) therapeutic benefits of vaccines on the burden of the pandemic and the time-to-elimination. The vaccination rates *ξ*_*v*_ = 7.4 × 10^*−*4^, 1.5 × 10^*−*3^, 3.0 × 10^*−*3^, *per* day correspond, respectively, to vaccinating approximately 2.5 × 10^5^, 5.0 × 10^5^, 1.0 × 10^6^ people *per* day. The vaccine-induced and natural immunity waning rate parameters, *ω*_*v*_ and *ω*_*r*_, are set to *ω*_*v*_ = 1*/*180 *per* day [40] and *ω*_*r*_ = 1*/*270 *per* day [42], respectively. The effect of therapeutic benefits of the vaccine depicted in (c) is incorporated by reducing the baseline values of the parameters (from Tables 3-4) that are related to development of severe disease (*r*), hospitalization (*ϕ*_*jI*_, *j* = 1, 2) and mortality (*δ*_*jI*_, *δ*_*jH*_, *j* = 1, 2) by 5%, in addition to increasing the baseline value of the parameters related to the recovery rate (*γ*_*jI*_, *γ*_*jA*_, *γ*_*jH*_, *j* = 1, 2) by 5%. The values of the other parameters of the model used in the simulation are as given in Tables 3-4.

#### 4.5.2 Mask fatigue and relaxation of mask mandates for fully-vaccinated individuals

To incorporate the effect of mask fatigue, or relaxation of mask mandates [43], in fully-vaccinated individuals into the model (2.5)-(2.6), we consider the *worst-case* scenario where all fully-vaccinated individuals opt to give up masking in public. To account for the worst case scenario of this (i.e., the case in which every fully-vaccinated individual abandons masking) in the model, we remove the state variable *S*_2*v*_, for the vaccinated susceptible individuals in the mask-wearing group 2, from the model. Further, we re-direct all the new vaccinated individuals from group 2 into the vaccinated class of the non-masking group 1 (i.e., we add the term *ξ*_*v*_*S*_2*u*_ from the equation for the rate of change of the *S*_2*u*_ population to that for the rate of change of the *S*_1*v*_ population, and the equation for *S*_2*v*_ is removed from the model) and also remove the term −*α*_12_*S*_1*v*_ from the equation for the rate of change of the *S*_1*v*_ population (to ensure that vaccinated individuals in group 1 do not move to the mask-wearing group 2). Simulations of the model (2.5)-(2.6), under this setting (and using the parameter values in Tables 3 and 4), depicted in Figure 8(b), show a marginal change in the peak number of new cases and the time-to-elimination, in comparison to the case when fully-vaccinated individuals do not completely give up masking (i.e., compare Figure 8(b) with Figure 7(a)).

#### 4.5.3 Therapeutic benefits of COVID-19 vaccines

Result from recent clinical trials have shown very promising therapeutic benefits for both the Pfizer and Moderna vaccines [15, 16]. In this section, we seek to use the multi-group model (2.5)-(2.6) to assess the impact of such benefits on the dynamics of the disease in the US. Since the model does not explicitly stratify the population of infected individuals according to whether they are vaccinated or not, a number of factors will come into play when estimating the overall impact of the therapeutic benefits, such as the high efficacy of the two vaccines (approximately 95%, thereby significantly reducing the size of breakthrough infections), level of vaccine hesitancy in the community and the current daily infection rate in the community. Taking all these into account, we consider it plausible, as a first approximation, to estimate the overall therapeutic benefits in the US, at the beginning of the third wave (characterized by low vaccination coverage (December 2020 until about February 2021), high disease burden (skyrocketing number of reported confirmed cases, hospitalizations and COVID-19 mortality), by a 5% reduction in severe or symptomatic illness, breakthrough transmission, hospitalization, and mortality, as well as a 5% increase in the rate of recovery from infection for vaccinated infected individuals. In other words, the effect of therapeutic benefits of the vaccine is incorporated into our model by reducing the baseline values of the parameters related to development of severe disease (*r*), hospitalization (*ϕ*_*jI*_, *j* = 1, 2) and mortality *δ*_*jI*_ and *δ*_*jH*_ with *j* = 1, 2) by 5%, in addition to increasing the baseline value of the parameter related to the recovery rate (*γ*_*jI*_, *γ*_*jA*_ and *γ*_*jH*_, with *j* = 1, 2). The simulation results obtained, for this hypothetical scenario, show a marked reduction in disease burden and a decrease in time-to-elimination (8(c)), in comparison to the case where such therapeutic benefits are not accounted for (Figure 7(a)). In particular, if one million people are vaccinated daily (i.e., if the vaccination rate is set at *ξ*_*v*_ = 3.0 × 10^*−*3^ *per* day), up to 37% decrease in the peak number of new cases could be achieved. Further, the time-to-elimination decreases by 17 days (compare green curves in Figures 7(a) and 8(a)). Higher reductions in disease burden, and more accelerated time-to-elimination, will be achieved if higher percentages of therapeutic benefits are assumed. It should be mentioned that a more rigorous way to introduce the impact of therapeutic benefits into the multi-group model (2.5)-(2.6) will be to further restructure the infected compartments of the model in terms of whether they are vaccinated or not (doing so will result in a model with at least 28 nonlinear differential equations, which may be difficult to track mathematically and statistically).

In summary, it is shown in this section (based on the parameter values used in our simulations) that, while waning natural and vaccine-derived immunity generally induces a relatively small increase in the burden of the pandemic, together with a correspondingly marginal increase in the time-to-elimination (in comparison to the case when these effects are not incorporated into the model), the therapeutic benefits of the vaccines offer a dramatic impact on the trajectory of the disease (by significantly reducing both the burden and time-to-elimination of the pandemic, in comparison to the case when such benefits are not accounted for in the model). Finally, it is worth stating that, although the simulations carried out in Section 4.5 are for the Pfizer and Moderna COVID-19 vaccines only (illustrated in Figure 7), similar simulations can also be carried out for the AstraZeneca and other vaccines with lower preventive effective efficacies. These simulations will, of course, show higher disease burden (owing to their reduced efficacy), in comparison to the case when Pfizer and Moderna vaccines are used.

## 5 Discussion and Conclusions

Since its emergence late in December of 2019, the novel Coronavirus pandemic continues to inflict devastating public health and economic burden across the world. As of January 24, 2021, the pandemic accounted for over 100 million confirmed cases and 2.1 million fatalities globally (the United States accounted for 25, 123, 857 confirmed cases and 419, 204 deaths). Although control efforts against the pandemic have focused on the use of non-pharmaceutical interventions, such as social-distancing, face mask usage, quarantine, self-isolation, contacttracing, community lockdowns, etc., a number of highly-efficacious and safe anti-COVID-19 vaccines have been developed and approved for use in humans. In particular, the two FDA-approved vaccines (manufactured by Moderna Inc. and Pfizer Inc.) have estimated protective efficacy of about 95%. Furthermore, AstraZeneca vaccine, developed by the pharmaceutical giant, AstraZeneca and University of Oxford (which is currently being used in the UK and other countries) has protective efficacy of 70%. Mathematics (modeling, analysis and data analytics) has historically been used to provide robust insight into the transmission dynamics and control of infectious diseases, dating back to the pioneering works of the likes of Daniel Bernoulli in the 1760s (on smallpox immunization), Sir Ronald Ross and George Macdonald between the 1920s and 1950s (on malaria modeling) and the compartmental modeling framework developed by Kermack and McKendrick in the 1920s [45–47]. In this study, we used mathematical modeling approaches, coupled with rigorous analysis, to assess the potential population-level impact of the use of the three currently-available vaccines in curtailing the burden of the COVID-19 pandemic in the U.S. We have also assessed the impact of other non-pharmaceutical interventions, such as face mask and social-distancing, implemented singly or in combination with any of the three vaccines, on the dynamics and control of the pandemic.

We developed a novel mathematical model, which stratifies the total population into two subgroups of individuals who habitually wear face masks in public and those who do not. The resulting two group COVID-19 vaccination model, which takes the form of a deterministic system of nonlinear ordinary differential equations, was initially fitted using observed cumulative COVID-induced mortality data for the U.S. Specifically, we fitted the model with the cumulative data corresponding to the period when the U.S. was experiencing the third wave of the COVID-19 pandemic (estimated to have started around October 12, 2020). In addition to allowing for the assessment of the population-level of each of the three currently-available vaccines, the model also allows for the assessment of the initial size of the population of individuals who habitually wear face masks in public, as well as assessing the impact of increase in the baseline social-distancing compliance attained as of October 12, 2020. After the successful calibration of the model, we carried out rigorous asymptotic stability analysis to gain insight into the main qualitative features of the model. In particular, we showed that the disease-free equilibrium of the model is locally-asymptotically stable whenever a certain epidemiological threshold, known as the *control reproduction number* (denoted by ℛ_*c*_), is less than unity. The implication of this result is that (for the case when ℛ_*c*_ *<* 1), a small influx of COVID-infected individuals will not generate an outbreak in the community.

The expression for the reproduction number (ℛ_*c*_) was used to compute the nationwide vaccine-induced *herd immunity* threshold for a special case of the model where change of masking behavior is not allowed. The herd immunity threshold represents the minimum proportion of the susceptible U.S. population that needs to be vaccinated to ensure elimination of the pandemic. Simulations of our model showed, for the current baseline level of social-distancing in the U.S. (and baseline level of initial size of the population of face masks wearers), herd immunity can be achieved in the U.S. using the AstraZeneca vaccine if at least 80% of the susceptible population is vaccinated. The threshold herd immunity level needed when either the Pfizer or Moderna vaccine is used reduces to 59%. Our simulations further showed that the level of herd immunity needed to eliminate the pandemic decreases, for each of the three currently-available vaccines, with increasing levels of baseline social-distancing compliance. In particular, the baseline social-distancing achieved at the beginning of our simulation period (i.e., the level of social-distancing in the U.S. as of October 12, 2020) is increased by 10%, the herd immunity requirement for the AstraZeneca or Pfizer/Moderna vaccine reduced, respectively, to 73% and 54%. Furthermore, if the baseline social-distancing is increased by 30%, the herd immunity threshold needed to eliminate the pandemic using the AstraZeneca or Pfizer/Moderna vaccine reduced to a mere 53% and 39%, respectively. In other words, this study showed that the prospect of achieving vaccine-derived herd immunity, using any of the three currently-available vaccines, is very promising, particularly if the vaccination program is complemented with increased levels of baseline social-distancing.

The multigroup nature of the model we developed in this study, where the total population is stratified into the two groups of those who habitually face mask in public and those who do not (with back-and-forth transitions between the two groups allowed), enabled us to assess the population-level impact of the initial sizes of the two groups in curtailing the spread of the pandemic in the United States. We assessed this by simulating the model during the beginning of the third wave of the pandemic in the U.S. (starting from October 12, 2020), and used the proportion of masks-wearers embedded in the cumulative mortality data we used to fit the model as the baseline. Our study emphasized the fact that early adoption of mask mandate plays a major role in effectively reducing the burden (as measured in terms of cumulative mortality) of the pandemic. This effect is particularly more pronounced when individuals in the face masks-wearing group do not change their behavior and transition to the non-mask wearing group (and non-mask wearers adopt a masks-wearing habit). Our study further showed that, for the case where the aforementioned back-and-forth transitions between the masks-wearing and the non-mask wearing groups is allowed, there is a threshold level of the initial size of the proportion of face masks-wearers above which the disease burden will be reduced, below which the disease burden actually increases. Our study estimated this threshold value of the initial size of the masks-wearing group to be about 50%. The epidemiological implication of this result is that the continued implementation of face masks use strategy (particularly at the high initial coverage level) will be highly beneficial in effectively curtailing the pandemic burden between now and the time when the two FDA-approved vaccines become widely available to the general public in the U.S. (expected to be around mid March to mid April of 2021).

We further showed that the time-to-elimination of COVID-19 in the U.S., using a vaccine (and a non-pharmaceutical intervention), depended on the daily vaccination rate (i.e., number of people vaccinated *per* day) and the level of increase in baseline social-distancing compliance achieved at the onset of the third wave of the pandemic (October 12, 2020). Specifically, our study showed that the COVID-19 pandemic can be eliminated in the U.S. by early May of 2021 if we are able to achieve moderate level of daily vaccination rate (such as vaccinating 1 million people every day) and the baseline social-distancing compliance achieved on October 12, 2020 is increased by 10% (and sustained). It should, however, be mentioned that the time-to-elimination is sensitive to the level of community transmission of COVID-19 in the population (it is also sensitive to the effectiveness and coverage (compliance) levels of the other (non-pharmaceutical) interventions, particularly face mask usage and social-distancing compliance, implemented in the community). Specifically, our study was carried out during the months of November and December of 2020, when the United States was experiencing a devastating third wave of the COVID-19 pandemic (recording on average over 200, 000 confirmed cases *per* day, together with record numbers of hospitalizations and COVID-induced mortality). This explains the somewhat *longer* estimated time-to-elimination of the pandemic, using any of the three currently-available vaccines, for the case where social-distancing compliance is kept at the baseline level. The estimate for the time-to-elimination (using any of the three currently-available vaccines) will be shorter if the community transmission is significantly reduced (as will be vividly evident from the reduced values of the transmission-related and mortality-related parameters of the re-calibrated version of our model).

It is worth emphasizing that at the time this study was carried out (December 2020), it was unclear whether natural or vaccine-induced immunity to COVID-19 waned over time. It was also unclear whether the then new vaccines that received FDA’s Emergency Use Authorization (Pfizer and Moderna) offer therapeutic benefits (such as reducing severe disease, hospitalization and deaths, in addition to accelerating recovery rate in vaccinated infected individuals). However, by the time we are reviewing the manuscript (June 2021), new data and studies have provided clarity on waning immunity to COVID-19 [41, 42] and on the therapeutic benefits of some of the COVID-19 vaccines [15, 16]. Furthermore, the U.S. Centers for Disease Control and Prevention has modified its guidelines on masking, allowing fully-vaccinated individuals not to wear masks under certain circumstances [43]. Consequently, we adapted the multi-group model we developed to allow for the assessment of the aforementioned new facts associated with the COVID-19 dynamics. Specifically, we adapted and simulated the model to assess the impact of waning immunity (both natural and vaccine-derived), mask fatigue and relaxation of mask mandates for fully-vaccinated individuals and the therapeutic benefits of the FDA-authorized vaccines on the disease burden (measured in terms of peak daily cases) and time-to-elimination of the pandemic in the US. The simulations were carried out for the hypothetical scenario that the vaccination program was started at the beginning of the third wave of the pandemic in the US (i.e., in October of 2020). Since the vaccines were not available until December 2020, and large scale vaccine rollout was only achieved some time in end of March 2021 or early April 2021, we adapted our conclusions appropriately to account for this time lag. Consequently, our simulation results, for these settings, show that, while waning natural and vaccine-derived immunity induces only a relatively marginal increase in both the burden and time-to-elimination of the pandemic, incorporating therapeutic benefits of the vaccine into the model causes a dramatic reduction in both the burden and time-to-elimination. If the impacts of therapeutic benefits are incorporated into the model from the very beginning of the third wave of the pandemic (October 2020), our simulations show that the pandemic could theoretically be eliminated in the US by as early as late May, 2021 (note that, in the absence of such therapeutic benefits, our study estimated the time-to-elimination to be some time in October, 2021).

In summary, our study suggest that the prospects of COVID-19 elimination in the U.S. is very promising, using any of the three currently-available vaccines. The elimination prospects are greatly enhanced if the therapeutic benefits of the approved vaccines are incorporated into the multi-group model we developed and used in this study. While, for the baseline scenario, the AstraZeneca vaccine requires at least 80% of the US population to be vaccinated to achieve herd immunity (needed for the elimination of the pandemic), such herd immunity can be achieved using any of the two FDA-approved vaccines (Pfizer or Moderna) if only 59% of Americans are vaccinated. The prospects of eliminating COVID-19 using any of the three vaccines is greatly enhanced if the vaccination program is combined with a social-distancing strategy that increases the baseline compliance level of the social-distancing attained during the beginning of the third wave of the COVID-19 pandemic in the United States. In fact, our simulations strongly suggest that COVID-19 can be eliminated in the U.S. in 2021, and as early as May 2021, depending on the level of increase in baseline social-distancing compliance. In other words, if we can continue to maintain social-distancing, while large scale vaccination is being implemented, COVID-19 can be history…and life can begin to return to normalcy or near-normalcy, in the spring or fall of 2021.

Some of the limitations of our model include not explicitly accounting for some important heterogeneities, such as age-structure, risk-structure, and vaccine dose structure, and the impacts of SARS-CoV-2 variants. Accounting for these may alter our results, especially during the early days of the vaccine administration (e.g., from December 2020 to April 2021) when the vaccine doses were generally in limited supply and needed to be prioritized to high-risk groups. Hence, our simulation results and conclusions should be interpreted with these limitations in mind. Further, while our multi-group model did not explicitly account for some other factors potentially relevant to COVID-19 dynamics, such as waning of natural and vaccine-derived immunity to COVID-19, mask fatigue and relaxation of mask mandates for fully-vaccinated individuals and the impacts of therapeutic benefits of the approved vaccines, our multi-group model was robust enough to allow for the assessment of these factors. We showed that, while incorporating waning of immunity and mask fatigue and relaxation of mask mandates in fully-vaccinated individuals in the model we developed only caused a marginal increase in disease burden and time-to-elimination, incorporating the impacts of the therapeutic benefits of the approved vaccines (even at a relatively low overall rate) resulted in a dramatic reduction in both the disease burden and time-to-elimination of the pandemic.

## Data Availability

All data used for this study is freely available from public sources identified in the manuscript.

## Acknowledgments

One of the authors (ABG) acknowledge the support, in part, of the Simons Foundation (Award #585022) and the National Science Foundation (Grant Number: DMS-2052363). CNN acknowledges the support of the Simons Foundation (Award #627346). GAN acknowledge the grants and support of the Cameroon Ministry of Higher Education, through the initiative for the modernisation of research in Higher Education. All authors wish to express their deepest sympathy to the families of the victims of the SARS-CoV-2, and extend profound appreciation to the frontline healthcare workers for their heroic effort and sacrifices to save the lives of others. The authors are very grateful to the two anonymous reviewers for their very constructive comments, which have significantly enhanced the quality and clarity of the manuscript.

## Appendix I: Entries of the Non-negative Matrix *F*

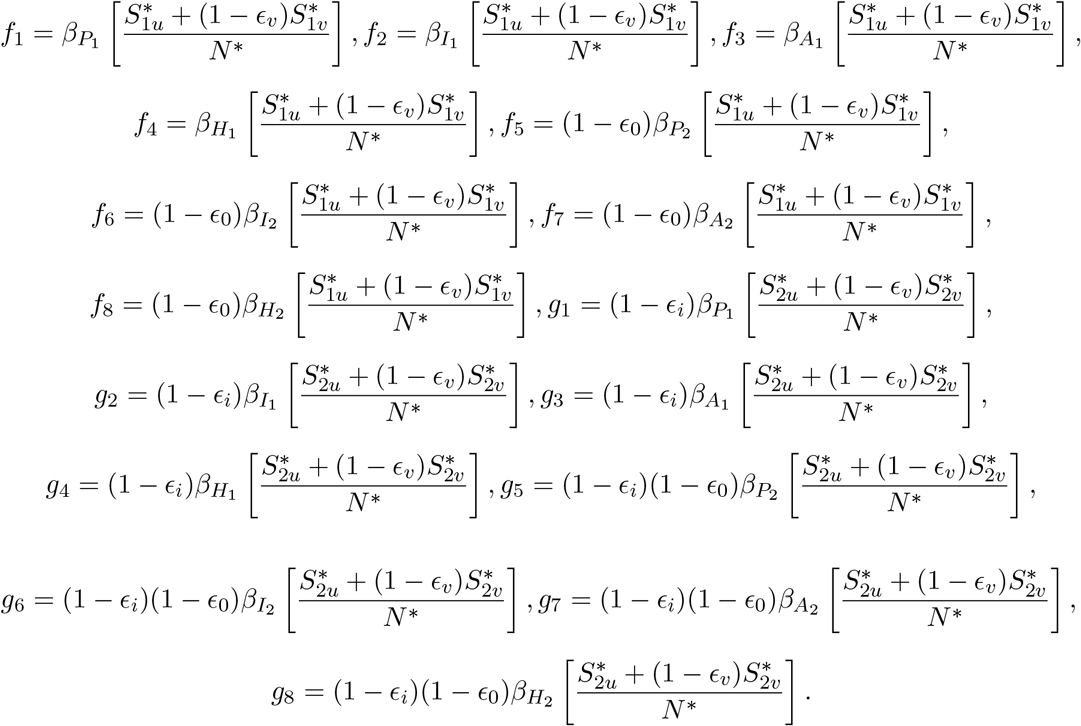

## Notes

### Competing Interest Statement

The authors have declared no competing interest.

### Summary of Updates

We have added a new section (Section 4.5), where we describe how the model can be extended to incorporate waning immunity, mask fatigue among fully-vaccinated individuals, and the therapeutic benefits of the vaccines. Also, we have added Figure 8 to illustrate the impact of these modifications. Furthermore, a paragraph on the limitations of the study has been added in the discussion section.

